# Fatal cytokine release syndrome by an aberrant FLIP/STAT3 axis

**DOI:** 10.1101/2021.05.04.21256298

**Authors:** Chiara Musiu, Simone Caligola, Alessandra Fiore, Alessia Lamolinara, Cristina Frusteri, Francesco Domenico Del Pizzo, Francesco De Sanctis, Stefania Canè, Annalisa Adamo, Francesca Hofer, Roza Maria Barouni, Andrea Grilli, Serena Zilio, Paolo Serafini, Evelina Tacconelli, Katia Donadello, Leonardo Gottin, Enrico Polati, Domenico Girelli, Ildo Polidoro, Piera Amelia Iezzi, Domenico Angelucci, Andrea Capece, Ying Chen, Zheng-Li Shi, Peter J. Murray, Marco Chilosi, Ido Amit, Silvio Bicciato, Manuela Iezzi, Vincenzo Bronte, Stefano Ugel

**Author notes:** shared first authors (Chiara Musiu, Simone Caligola, Alessandra Fiore). shared last authors (Vincenzo Bronte, Stefano Ugel).

## Abstract

Inflammatory responses rapidly detect pathogen invasion and mount a regulated reaction. However, dysregulated anti-pathogen immune responses can provoke life-threatening inflammatory pathologies collectively known as cytokine release syndrome (CRS), exemplified by key clinical phenotypes unearthed during the SARS-Cov-2 pandemic. The underlying pathophysiology of CRS remains elusive. We found that FLIP, a protein that controls caspase-8 death pathways, was highly expressed in myeloid cells of COVID-19 lungs. FLIP controlled CRS by fueling a STAT3-dependent inflammatory program. Indeed, constitutive expression of a viral FLIP homologue in myeloid cells triggered a STAT3-linked, progressive and fatal inflammatory syndrome in mice, characterized by elevated cytokine output, lymphopenia, lung injury and multiple organ dysfunctions that mimicked human CRS. As STAT3-targeting approaches relieved inflammation, immune disorders, and organ failures in these mice, targeted intervention towards this pathway could suppress the lethal CRS inflammatory state.

**One sentence summary:** FLIP-expressing myeloid cells are key drivers of CRS through aberrant overexpression of STAT3 pathway. STAT3-targeting is effective in mitigating CRS like severe COVID-19.

## Introduction

Host responses to pathogens are ordered, time-dependent and tissue-compartmentalized, coordinated by the release of soluble factors, such as growth factors and inflammatory cytokines, which engage, activate and regulate innate immune cells(*1*). Since the magnitude of the immune response is generally consistent with the pathogen load and restrained to the invasion area, cytokines with short half-life have a limited action at sites of inflammation and favor the local activation of immune cells. However, persistent infections or an uncontrolled microbial burden can prompt higher output of cytokines, which fuel emergency hematopoiesis to mobilize an increased number of leukocytes from the bone marrow and thus counterbalance the myeloid cell depletion in periphery(*2*). Under these circumstances, the increase in cytokines beyond the normal thresholds can cause a catastrophic cytokine storm known as cytokine release syndrome (CRS), which eventually leads to multiple organ failures(*3*). This systemic pathology is not only associated with disseminated bacterial or viral infections(*4, 5*) but also induced by cancer immunotherapy, i.e. chimeric antigen receptor (CAR) T cells infusion or antibody-based therapy(*6-10*), stem cell transplantation settings(*11*), as well as linked to either autoimmune or genetic disorders(*12, 13*). Patients with CRS frequently display respiratory symptoms including tachypnea that progress to acute respiratory distress syndrome (ARDS)(*14*), severe kidney injury, hepatobiliary damage, cardiomyopathy and neurological dysfunctions(*3*).

Accumulating evidence suggests a close relationship between CRS and the pandemic of coronavirus disease 2019 (COVID-19) induced by severe acute respiratory syndrome coronavirus 2 (SARS-CoV-2)(*15-17*). Indeed, pathophysiological features of severe COVID-19 patients were often associated with pulmonary involvement that can require invasive mechanical ventilation in intensive care units (ICU)(*18-21*). After virus entry, SARS-CoV-2 induces endothelial cell damage, complement activation, thrombin production and fibrinolysis inhibition that result in pulmonary intravascular coagulation, vascular microthrombi formation and, ultimately, severe vasculopathy, acute myocardial infarction and stroke(*22, 23*). Moreover, several studies have reported SARS-CoV-2 organotropism in myocardial, renal, neural and gastrointestinal tissues, confirming COVID-19 as a complex pathology with multiple manifestations(*24*). These multi-organ damages may be associated to an either direct viral toxicity or virus-dependent dysregulation of the immune system(*25-30*). Thus, activation of innate immune system can be considered the main hallmark of SARS-CoV-2-associated CRS, as recently reviewed(*31*). Indeed, myeloid cells are actively involved in the establishment of acute lymphopenia, microvascular dysfunctions and organ failure, all key features of COVID-19-associated CRS(*27, 32-34*). To date, however, a molecular understanding of the SARS-CoV-2-dependent myeloid cell reprogramming remains elusive.

We previously identified the anti-apoptotic cellular and viral FLICE (FADD-like IL-1β-converting enzyme)-like inhibitory proteins (hereafter c-FLIP and v-FLIP respectively) as a factor that “reprogrammed” monocytes(*35*). FLIP isoforms are conventionally described to control cell survival and proliferation as caspase-8 inhibitor and/or NF-κB activator(*36-40*). However, FLIPs regulate different biological processes based on their protein structure (e.g. the presence of death effector domains in tandem)(*41*) or cellular localization (e.g. trafficking between the nucleus and cytoplasm)(*42, 43*). The up-regulation of FLIP proteins in monocytes promoted the acquisition of unconventional phenotype characterized by the concurrent expression of immunosuppressive (e.g. programmed death-ligand 1 (PD-L1), IL-10) and pro-inflammatory (e.g. interleukin (IL)-1β, IL-6, tumor necrosis factor (TNF)-α) features, which partially depend on the nuclear translocation of the complex FLIP/nuclear factor kappa B p50 (NF-κB p50) protein(*35*). In transgenic ROSA26.vFLIP;LyzM-CRE mice, where Kaposi’s sarcoma virus vFLIP expression is enforced in myeloid cells, a severe and lethal inflammatory pathology developed, resembling Kaposi sarcoma-associated herpesvirus inflammatory cytokine syndrome(*35*).

We suspected FLIP- and pSTAT3-expressing myeloid cells were linked to COVID-19-associated CRS since their accumulation was shared by both human (h)ACE2-expressing transgenic mice and patients infected by SARS-CoV-2. Moreover, monocytes isolated from COVID-19 patients showed high myeloid expression of c-FLIP and pSTAT3 that correlated with their immunosuppressive properties. We established that vFLIP transgenic mice mirror both the immune dysfunctions and the bronchoalveolar immune landscape of patients affected by severe COVID-19. We used this unique model to assess both systemic and myeloid-targeted STAT3 interference approaches to resolve uncontrolled inflammation and acute disease manifestations.

## Results

### SARS-CoV-2 infection induced c-FLIP overexpression in both COVID-19 patients and virus-infected, HFH4-hACE2 transgenic mice

Viruses have evolved a myriad of ways to escape host apoptotic process and thereby preserve infected cells from early death, which eliminates the replicative niche(*44*). Several viruses including hepatitis C virus(*45*), hepatitis B virus(*46*), human T cell leukemia virus-1(*47*), human immunodeficiency virus 1(*48*), Epstein Barr Virus(*49*) and influenza A virus(*50*) induce the anti-apoptotic protein c-FLIP, which blocks caspase-mediated cell death(*40*). Poxviruses and herpesviruses encode proteins with high homology to c-FLIP, such as the Kaposi’s herpesvirus K13/vFLIP and the herpesvirus saimiri orf71, which harbor DED domains responsible for blocking procaspase cleavage, preventing apoptosis and favoring viral latency(*36*). Thus, FLIP expression is linked to viral replication by suppressing host cell death.

To examine c-FLIP alterations in COVID-19, we analyzed lung autopsy samples from patients infected by SARS-CoV-2 (COVID-19; n=23) or affected by bacterial pneumonia (BP; n= 4) or other diseases (NRD; n = 4) (Supplementary information, Table S1a). In the latter group, the respiratory tract was not altered by severe inflammation and showed a tissue structure almost devoid of pathological aspects (Fig. 1a, first line). By contrast, the histopathological features of the lungs from COVID-19 patients were showed heterogeneous inflammation and tissue damage (Supplementary information, Fig. S1a)(*22, 51*). When we evaluated the number of CD68^+^ myeloid cells, which encompass alveolar macrophages, monocytes/interstitial macrophages and histiocytes(*52, 53*), for the expression of FLIP, COVID-19 samples displayed a variable number of FLIP-expressing CD68^+^ cells (Fig. 1a, second line) suggesting an accumulation of FLIP^+^ myeloid cells during COVID-19 progression. Previously, we found enforced FLIP expression reprogramming myeloid cells into immune regulatory elements through the aberrant activation of several molecular pathways, including STAT3-dependent signaling(*35*). Since STAT3 hyperactivation was advanced as the orchestrator of most commonly COVID-19-associated features, such as rapid coagulopathy, thrombosis, tissue fibrosis, production of inflammatory cytokines and chemokines, as well as T cell lymphopenia, we next evaluated the expression of pSTAT3 in the selected pathological lung fields. We detected a weak pSTAT3 expression in NRD samples, while a limited pSTAT3 pattern was restricted to stromal cells in bacterial pneumonia sections. However, consistent and diffuse expression of pSTAT3 was shared in COVID-19 samples by numerous cell types; among them, several CD68^+^FLIP^+^ alveolar macrophages (Fig. 1a, third and fourth lines), histiocytic cells (Fig. 1a, fifth and sixth lines) and monocytes/interstitial macrophages (Supplementary information, Fig. S1b). Interestingly, CD68^+^FLIP^+^pSTAT3^+^ cells were present in 56.5% (13/23) of analyzed cases and their presence significantly correlated with a shorter time to fatal evolution, expressed as number of hospitalization days (i.e. absence of CD68^+^FLIP^+^pSTAT3^+^ cells (n=10) 25 ± 14.4 days *vs*. presence of CD68^+^FLIP^+^pSTAT3^+^ cells (n=13) 18 ± 9.3 days; p=0.0223) (Supplementary information, Table S1b).

**Fig. 1.**
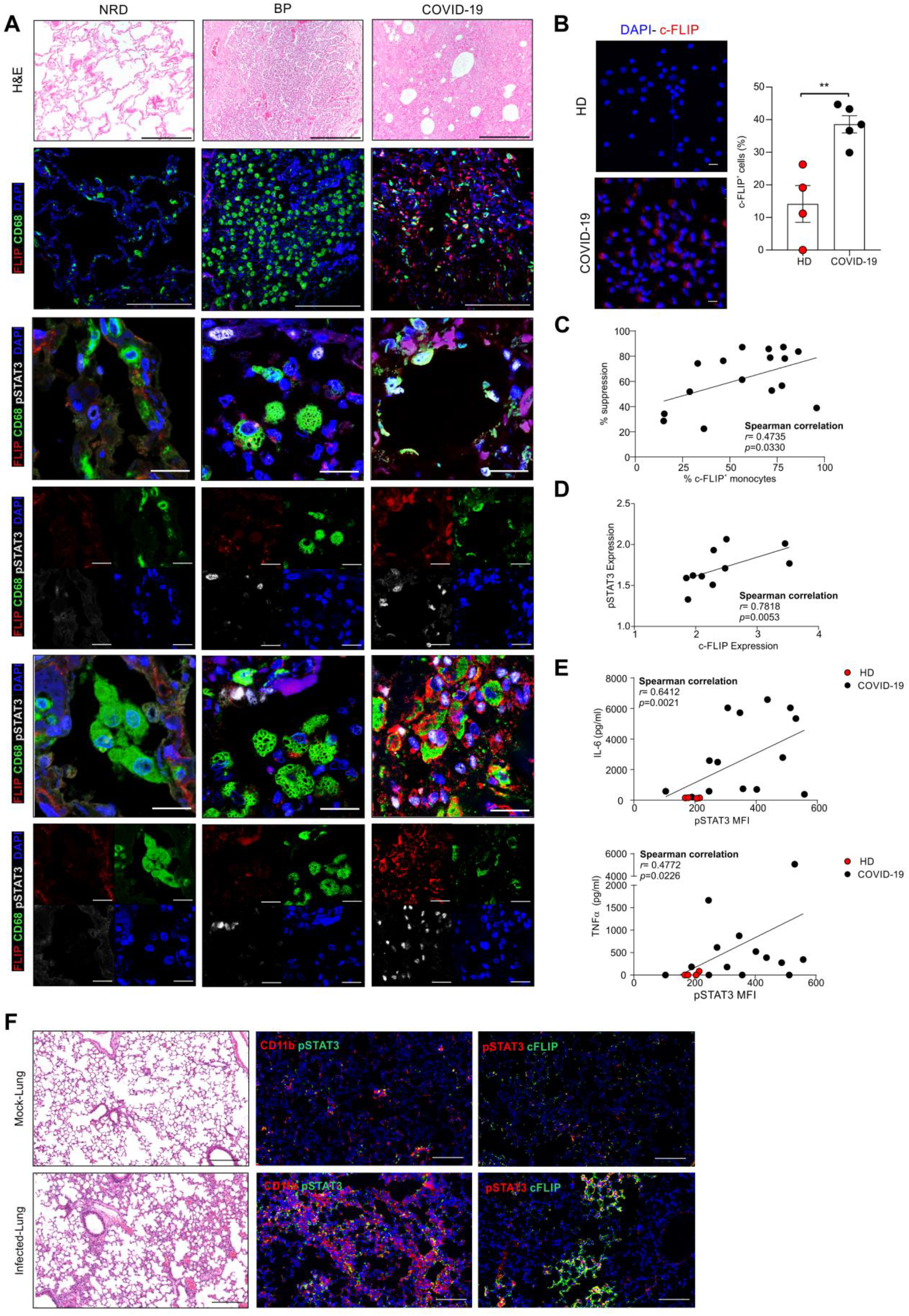
c-FLIP and pSTAT3 expression in SARS-CoV-2-infected hosts. **(A)** Representative H&E-stained microscopy images of lung tissue of non-respiratory disease (NDR), bacterial pneumonia (BD) and COVID-19 patients (upper panel). Scale bar, 200 μm. Representative immunofluorescence (IF) staining of alveolar macrophages (second, third and fourth lines) and histiocytic cells (fifth and sixth lines). Cells were stained for CD68 (green), c-FLIP (red), pSTAT3 (white) and nuclei (blue). Scale bar, 20 μm. **(B)** Quantification and representative IF staining of c-FLIP in monocytes purified from healthy donors (HD, n=4) or COVID-19 patients (n=5). Cells were stained for nuclei (blue) and c-FLIP (red). Scale bar, 10 μm. **(C)** Correlation between percentage of suppression and c-FLIP expression of circulating monocytes isolated from COVID-19 patients (n=16). **(D)** Correlation between pSTAT3 and c-FLIP expression in circulating CD14^+^ cells isolated from COVID-19 patients (n=10). **(E)** Correlation between the release of IL-6 or TNF-α cytokines and pSTAT3 expression in circulating CD14^+^ cells from HD (red, n=4) and COVID-19 patients (black, n=13). **(F)** Representative hematoxylin and eosin (H&E)-stained microscopy images of lung tissue of HFH4-hACE2 transgenic mice SARS-CoV-2-infected or mock-infected mice. Scale bar, 200 μm. pSTAT3, c-FLIP and CD11b expression levels were detected by indirect immunofluorescence (IFA) staining. Scale bar, 100 μm. Correlation analysis was performed by Spearman’s rank correlation (**C, D, E**).

We assessed c-FLIP and pSTAT3 expression in fresh circulating monocytes isolated from SARS-CoV-2 infected individuals (Supplementary information, Table S1c). Immunofluorescence analysis revealed an increased expression of c-FLIP in monocytes isolated from COVID-19 patients compared to heathy donors (HDs) (Fig. 1b), together with a linear correlation between c-FLIP-expression in monocytes and their immunosuppressive properties (Fig. 1c). PD-L1 was enhanced in c-FLIP-expressing CD14^+^ cells (Supplementary information, Fig. S1c), in agreement with our previous findings in pancreatic ductal adenocarcinoma (PDAC) patients(*35*), hinting to common mechanisms of innate immunity modulation in COVID-19 and cancer.

We next sought to determine whether c-FLIP-expressing monocytes exhibited concomitant activation of STAT3. We identified a significant direct correlation between pSTAT3 and c-FLIP expression in circulating CD14^+^ cells isolated from COVID-19 patients (Fig. 1d), hinting to the aberrant activation of FLIP/STAT3 axis in myeloid cells not only at pulmonary site but also in periphery. The STAT3 pathway in myeloid cells is relevant for acquiring immunosuppressive functions(*54-56*) and for driving production of cytokines during immune disorders(*57*), two conditions jointly cooperating to establish a severe lymphopenia, one the signs of clinical severity in COVID-19 patients. Monocytes isolated from COVID-19 patients secreted a greater amount of cytokines, on a per-cell basis, which correlated with the pSTAT3 expression (Fig. 1e and Supplementary information, Fig. S1e), consistent with published data about the monocyte contribution to the cytokine storm(*58, 59*).

To establish a direct link between FLIP and pSTAT3 dysregulation following SARS-Cov-2 infection, we analyzed the lung of mice transgenic for hACE2 (HFH4-hACE2 mice) that were intranasally infected with either SARS-CoV-2 or mock virus, as previously described(*60*). Examination of lung tissues 7 days after virus challenge demonstrated that SARS-CoV-2 infection induced severe pneumonia characterized by increased CD11b^+^ myeloid cell accumulation in perivascular and alveolar locations (Fig. 1f). Of note, lung-infiltrating myeloid cells in SARS-CoV-2-infected mice expressed higher p-STAT3 levels than the control group (Fig. 1f). Moreover, we found an increased expression of p-STAT3 in c-FLIP-expressing cells, which morphologically resemble myeloid cells, in the lung of SARS-CoV-2-infected mice (Fig. 1f).

Collectively, these data establish that myeloid subsets are converted into FLIP- and pSTAT3-expressing elements characterized by pro-inflammatory and immunosuppressive features in COVID-19.

### vFLIP overexpression in myeloid cell lineage induced pulmonary and systemic pathological features of CRS, including the fatal outcome

Since transgenic mice expressing v-FLIP in myeloid cells die prematurely within four weeks of life due to systemic immune disorders(*35*), we engrafted sub-lethally ablated, immunocompetent mouse recipients with bone marrow (BM) isolated from either ROSA26.vFLIP;LyzM-CRE (CD45.2) or wild type mice (CD45.1) mixed together at different ratios. All engrafted mice developed weight loss (Supplementary information, Fig. S2a-b), systemic lymphopenia and extensive accumulation of myeloid cells in the spleen, where a subversion of splenic architecture was marked (Supplementary information, Fig. S2c), as well as in several organs leading to the development of multi-organ injuries and areas of fibrosis (Supplementary information, Fig. S2d), including the lung. For subsequent analyses in this study we employed BM chimeras (hereafter defined vFLIP mice), generated by transplantation of a 1:1 ratio of vFLIP^+^ and WT BM cells.

Since lung inflammation is the principal cause of life-threatening respiratory syndrome in CRS, including severe forms of COVID-19(*20*), we assessed the lung histopathology in vFLIP mice. Lungs of vFLIP mice showed diffuse interstitial myeloid infiltrate and alveolar damage, characterized by areas of fibrosis, lung consolidation, multinucleated cell clusters and tissue regions with peribronchial and perivascular infiltrate with associated intravascular thrombi. Severe cases also showed infarctions and extensive fibrosis (Fig. 2a-b, Supplementary information, Fig. S3a). Consistent with COVID-19 CRS, we detected pSTAT3 in myeloid cells localized in vFLIP lung tissues compared to normal mice (Fig. 2c). Considering the role of an aberrant immune response in the pathogenesis of COVID-19 by promoting alveolar inflammation and hyper-coagulation state in lung vessels(*26, 61*), we evaluated the presence of the endothelial dysfunction marker p-selectin in the pulmonary environment of vFLIP mice. P-selectin is normally stored in Weibel-Palade bodies of endothelial cells. After tissue injury, it is exposed in the vascular lumen where it mediates the adhesion and activation of platelets and leukocytes(*62*). In vFLIP mice, p-selectin was strongly expressed on the luminal surface of inflamed vessels of large and small caliber, whereas it was not detectable in the lungs of WT mice (Fig. 2d).

**Fig. 2.**
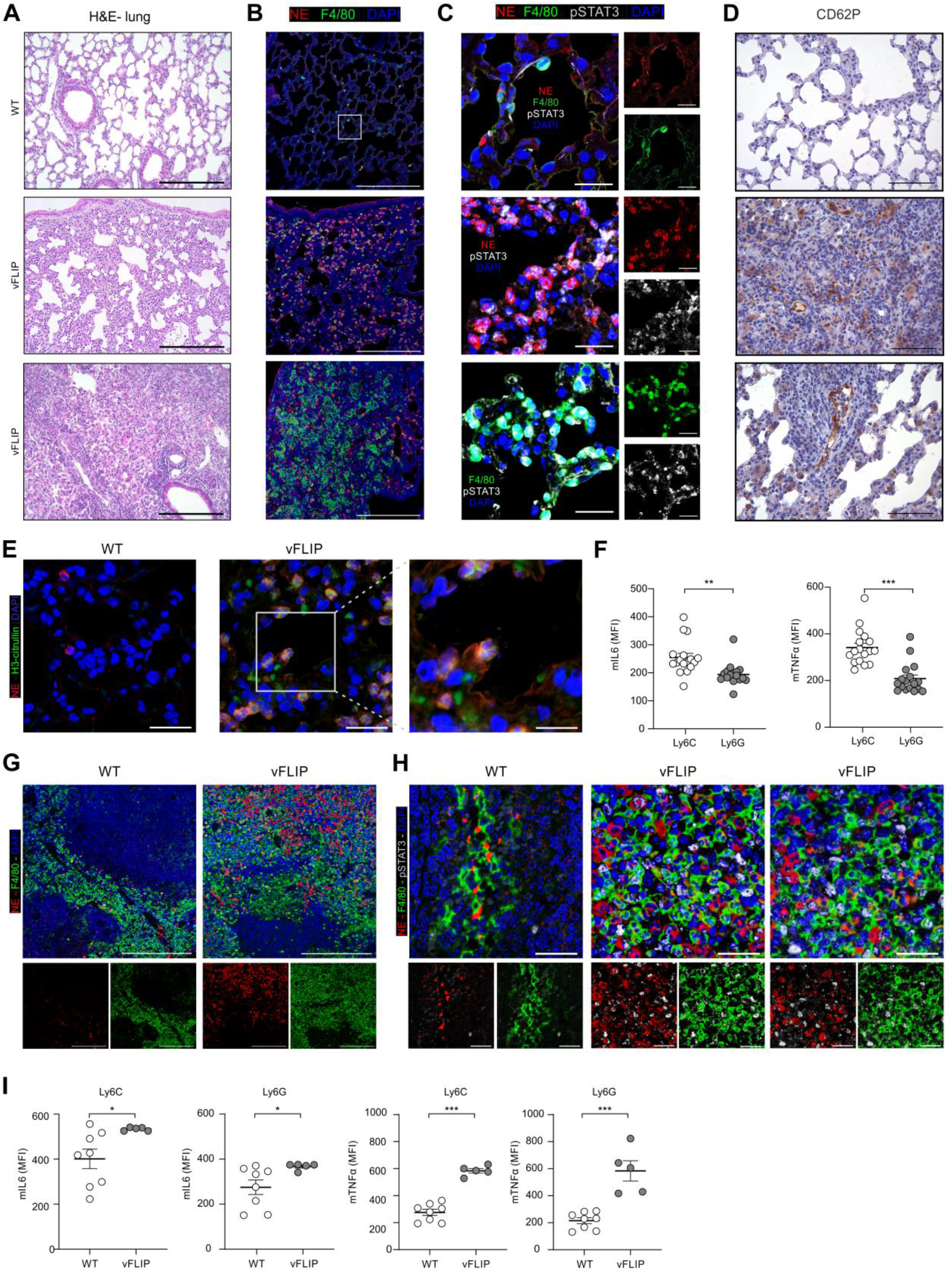
Local and systemic pSTAT3-dependent inflammation in vFLIP mice. **(A)** Representative H&E-stained microscopy images of lung of WT (upper panel) or vFLIP chimera mice. Scale bar, 200 μm. **(B)** Representative IF staining of lung-infiltrating neutrophils and macrophages in WT or vFLIP mice. Cells were stained for DAPI (blue), neutrophil elastase (NE) (red, middle panel) and F4/80 (green, bottom panel). Scale bar, 200 μm. **(C)** Representative IF staining of lung-infiltrating neutrophils and macrophages in WT or vFLIP mice. Scale bar, 20 μm. Cells were stained for DAPI (blue), NE (red, middle panel) or F4/80 (green, bottom panel) and pSTAT3 Tyr705 (grey). **(D)** CD62P presence in lung of WT (upper panel) or vFLIP mice by H&E staining. Scale bar, 100 μm. **(E)** Representative confocal analysis of NET in WT (left panel; 50 μm) or vFLIP mice (200 μm). Cells were stained for DAPI (white), NE (red) and H3cit (blue). **(F)** Dot plots of IL-6 and TNF-α in lung-infiltrating mononuclear (CD45.2^+^Ly6C^+^ cells) and polymorphonuclear (CD45.2^+^Ly6G^+^ cells) cells isolated from vFLIP mice (n=17). **(G)** Representative IF staining of spleen-infiltrating neutrophils (NE^+^ cells) and monocytes/macrophages (F4/80^+^ cells) in WT or vFLIP mice. Cells were stained for DAPI (blue), NE (red, middle panel) and F4/80 (green, bottom panel). Scale bar, 200 μm. **(H)** Representative IF staining of spleen-infiltrating neutrophils and macrophages in WT or vFLIP mice. Cells were stained for DAPI (blue), NE (red, middle panel) or F4/80 (green, bottom panel) and pSTAT3 Tyr705 (grey). **(I)** Dot plots of IL-6 and TNF-α in myeloid cells in spleen of WT (n=8) or vFLIP mice (n=9). Data are reported as mean ± S.E.M. *p ≤ 0.05, **p ≤ 0.01 and ***p ≤ 0.001 by Mann–Whitney test.

Among effector mechanisms of neutrophils during inflammatory processes, neutrophil-derived extracellular traps (NETs) have been linked with the pathology of lung damage in COVID-19(*61*). In line with this evidence, NETs, identified as extracellular DNA staining colocalizing with neutrophil elastase (NE) and citrullinated histone H3 (H3Cit) by confocal microscopy(*63*), were not found in lung of WT mice but present in parenchima and alveoli of v-FLIP mice (Fig. 2e). Since several stimuli trigger neutrophil activation and NETosis, including inflammatory cytokines and chemokines(*64, 65*), we evaluated whether myeloid cells could be a source of these soluble factors. By intracellular staining, we enumerated TNF-α- and IL-6-producing mononuclear (CD45.2^+^Ly6C^+^ cells) and polymorphonuclear (CD45.2^+^Ly6G^+^ cells) cells isolated from the lung of vFLIP mice. We could not isolate sufficient number of myeloid cells from WT mouse lungs. Inflammatory cytokine production was higher in monocytes than in neutrophils (Fig. 2f). Notably, a marked accumulation of myeloid cells was identified also in the spleen of vFLIP mice (Fig. 2g), in which high percentages of pSTAT3^+^ and inflammatory cytokine-producing monocytes were detected (Fig. 2h-i). We speculated that these cells might establish an unfavorable environment for T cells and, indeed, effector and helper T cells were heavily contracted while T regulatory lymphocytes (Tregs) significantly expanded in the spleen of vFLIP mice (Supplementary information, Fig. S3b-c), unveiling a pronounced systemic lymphopenia that mirrors COVID-19-associated CRS.

To gain broad insight into the immune landscape of CRS in vFLIP mice, we performed a single cell RNA sequencing (scRNA-seq) on lung-infiltrating cells. After preprocessing, integration and cell annotation (see Methods), a total of 31,274 mouse cell transcriptomes were obtained across WT and vFLIP mice. All the cells were visualized through t-distributed stochastic neighbor embedding (t-SNE) assessing the cell proportions across the different mouse samples to have a global overview of the cell composition (Supplementary information, Fig. S4a-b). When we compared the percentage of each cell type across WT and vFLIP conditions (Fig. 3a), vFLIP mice showed a higher proportion of neutrophils (∼65%) compared to WT mice (∼23%) and a marked decrease in T (∼5% vFLIP; ∼17% WT) and B lymphocytes (∼3% vFLIP; ∼21% WT), confirming the aforementioned histopatological data about pulmonary infiltration of neutrophils (Fig. 2c). Conversely, monocytes (∼9% vFLIP; ∼12% WT) and macrophages (∼13% vFLIP; ∼16% WT) were comparable between the two groups.

**Fig. 3.**
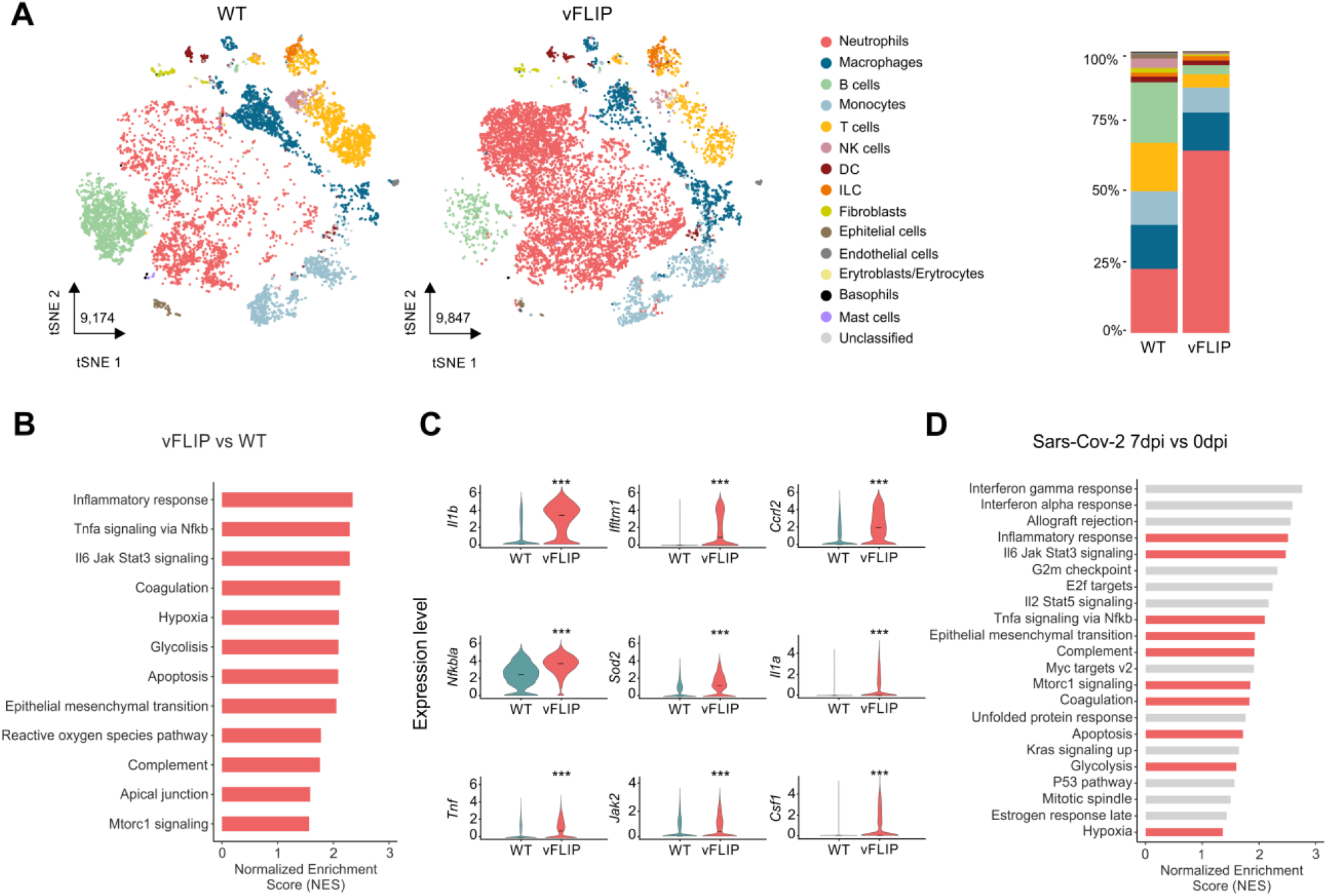
Lung immune landscape in vFLIP mice affected by cytokine release syndrome. **(A)** tSNE representation of scRNA-seq from 2 WT and 2 vFLIP mice samples (WT: 9,174; vFLIP: 9,847) colored according to cell type. Stacked bar plots representing cell type proportions across WT and vFLIP conditions. **(B)** Bar plot representing the up-regulated (NES > 0, adjusted p-value < 0.05) hallmark gene sets in the analysis of vFLIP vs WT cells obtained through GSEA analysis. **(C)** Violin plots showing the expression of key genes that drive the up-regulation of inflammatory response, TNF-α signaling via NF-κB and JAK-STAT3 signaling pathway in the lung of vFLIP mice. The asterisks denote statistically significant up-regulation in the comparison between vFLIP and WT conditions (*p < 0.05, **p < 0.01, ***p < 0.001). **(D)** Bar plot representing the up-regulated (NES > 0, adjusted p-value < 0.05) hallmark gene sets in bulk RNA-seq data obtained through GSEA analysis of ACE2-transgenic mice infected with Sars-CoV-2 (Winkler et al., 2020) comparing day 7 post infection (dpi) with mock-infected (0 dpi). Red bars refer to the gene sets enriched in both vFLIP and ACE2-transgenic mice.

To picture a global overview about biological pathways activated in vFLIP mice, we performed gene set enrichment analysis (GSEA) comparing vFLIP and WT cells. We noticed a significant up-regulation in inflammatory responses, TNF-α signaling via NF-κB and JAK-STAT3 signaling pathway that were among the top enriched gene sets (Fig. 3b). Notably, these processes were related to the over-expression of several pro-inflammatory mediators such as Il1b, Ccrl2, Il1a and Tnf (Fig. 3c). All these data are in line with results about pSTAT3 overexpression and TNF-α hyper-production in vFLIP mice (Fig. 2f), as well as previous data about FLIP controlling NF-κB(*35, 66*). To link our results from vFLIP mice with Sars-Cov-2 infection, we matched GSEA results obtained from vFLIP mice with GSEA analysis of bulk RNA-Seq data of lung from hACE2 transgenic mouse infected with Sars-Cov-2(*67*). Comparing vFLIP up-regulated pathways with those enriched following infection of hACE2 mice (day 7 post infection *vs* day 0, Fig. 3d), we found that SARS-CoV-2-induced inflammatory pathways were shared with vFLIP mice. Interestingly, these data are in line with transcriptomic analysis using additional animal models of SARS-CoV-2 infection(*68*).

Together, these results support the concept that CRS syndrome in vFLIP mouse model may mimic COVID-19 immunopathology.

### vFLIP mice and COVID-19 patients display similar inflammatory myeloid cell landscapes in pulmonary environment

To compare the pulmonary immune landscape of vFLIP mice and COVID-19 patients, we quantified the similarity between mouse lung-infiltrating leukocytes and 19,996 immune cells isolated from bronchoalveolar lavage fluids (BAL) of severe COVID-19 patients (n=7; WHO ordinal score 7) who were admitted to Intensive Care Units (ICU) of Verona Hospital(*32*). A procedure similar to mouse cell integration and annotation was performed in order to assess cell composition in human BAL samples (Supplementary information, Fig. S4c-d). In both mouse and human datasets, neutrophils and monocyte/macrophage populations were separated and re-integrated prior to clustering, in order to have a better resolution in subset identification and comparison.

Following clustering, mouse neutrophils comprised 4 subsets characterized by canonical neutrophil markers including S100-family genes and *Adam8* (cluster 0), inflammatory chemokines *Ccl3* and *Ccl4* (cluster 1), high expression of *Camp* and *Ngp* genes (cluster 2) and interferon response genes *Isg15/Isg20* (cluster 3) (Fig. 4a and Supplementary information, Table S2). Similarly, in COVID-19 BAL, we outlined 5 different clusters expressing marker genes similar to mouse neutrophil subsets. In particular, we observed clusters expressing canonical neutrophil markers such as *S100A8/S100A9* genes (cluster a), *CCL3/CCL4* chemokines as well as *CTSB* and *CSTB* genes (cluster b), interferon-response genes *IFIT1, IFIT2* and *ISG15/ISG20* (cluster c) and other 2 clusters expressing ribosomal (cluster d) and heat shock protein (HSPs) genes (cluster e) (Fig. 4b and Supplementary information, Table S2).

**Fig. 4.**
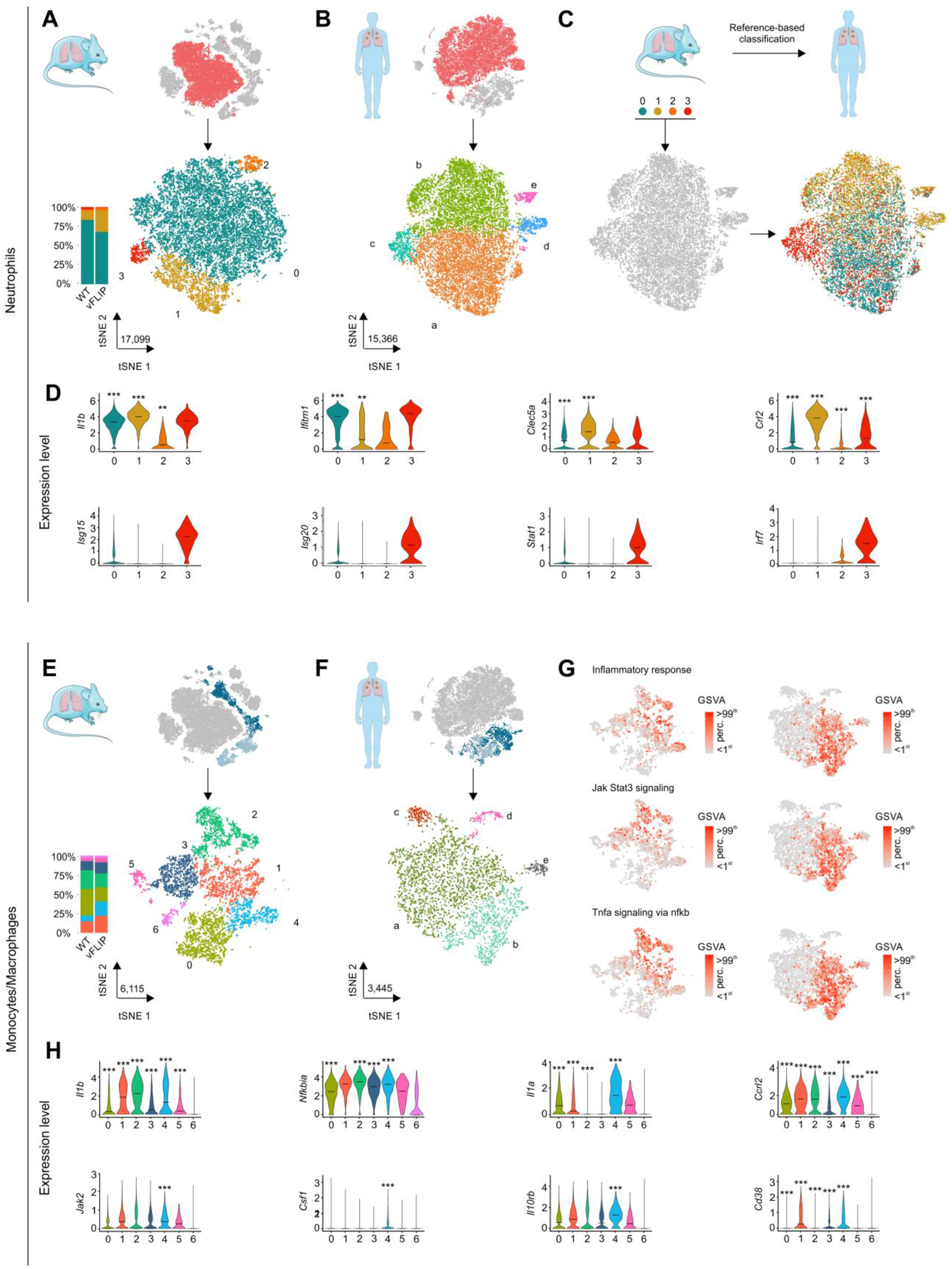
Unbiased comparison between lung-infiltrating cells of vFLIP mice and BAL-derived immune cells obtained from COVID-19 patients. **(A)** tSNE representation and stacked bar plot showing cluster analysis of neutrophils coming from scRNA-seq analysis of lung tissue of mice (n=7) and **(B)** COVID-19 patients BALs (n=7). Neutrophils of mouse (17,099) and human (15,366) are visualized through tSNE projection and colored according to cluster analysis. **(C)** Reference-based classification of BAL neutrophil clusters using average expression of mice neutrophil clusters (see Methods). **(D)** Violin plots showing the expression of key inflammatory and interferon response genes among mouse neutrophil clusters. The asterisks denote statistically significant up-regulation in the comparison between vFLIP and WT conditions (*p < 0.05, **p < 0.01, ***p < 0.001). **(E)** Subset analysis of monocytes/macrophages of mouse (n = 7) and **(F)** COVID-19 BALs (n = 7). Monocytes/macrophages of mouse (6,115) and human (3,445) are visualized through tSNE projection and colored according to cluster analysis. **(G)** GSVA scores for inflammatory response, JAK-STAT3 signaling and TNF-α signaling via NF-κB pathway across mouse (left) and BAL (right) monocytes/macrophage clusters. **(H)** Violin plots showing the expression level of key inflammatory response and JAK-STAT3 signaling pathway genes across mouse monocytes/macrophages subsets. The asterisks denote statistically significant up-regulation in the comparison between vFLIP and WT conditions (*p < 0.05, **p < 0.01, ***p < 0.001).

We used reference-based classification (see Methods section) to map mouse subsets into human subsets. The results confirmed a conserved structure among neutrophil clusters between the two species and pathologies (Fig. 4c). In fact, about 89% of the human cluster c was annotated as cluster 3 of mouse, about 81% of cluster d was annotated as cluster 1 and, finally, clusters a, b and e were mainly annotated as cluster 1 of mouse (60-65%). Conversely, cluster 2 appeared to be a mouse-specific neutrophil subset with no relevant correspondence in human subsets. These results recapitulate published reports (*69*), in which 3 conserved modules between mouse and human were characterized by the expression of *Ccl3/CCL3, Cstb/CSTB* (cluster 1 in vFLIP), type I interferon response genes such as *Ifit1/IFIT1, Irf7/IRF7* and *Rsad2/RSAD2* (cluster 3 in vFLIP), and neutrophils expressing canonical markers *S100a8-a9/S100A8-A9* (cluster 0 in vFLIP).

Even though mouse clusters 1 and 3 were not the most abundant neutrophil subsets (Fig. 4a) in vFLIP mice, they were the most dysregulated in terms of cell proportion compared to WT. Indeed, Cluster 1 displayed more than 2-fold increase (∼27% vFLIP; ∼11% WT) while cluster 3 about 3-fold decrease in vFLIP compared to WT (∼3% vFLIP; ∼1% WT). Furthermore, GSEA analysis at the cluster level revealed several leading genes involved in neutrophil inflammatory response in cluster 1 (Supplementary information, Table S2), such as *Il1b, Ifitm1, Clec5A* and *Ccrl2*, which were up-regulated in vFLIP compared to WT mice (Fig. 4d and Supplementary information, Table S2).

We detected 7 clusters in mouse mononuclear phagocyte compartment (monocytes/macrophages) (Fig. 4e). Among macrophages we could identify alveolar macrophages expressing *Mrc1, Krt79* and *Krt19* genes (cluster 0), macrophages expressing either inflammatory cytokines such as *Cxcl3, Cxcl1* and *Il1a* (cluster 4) or proliferation markers *Mki67* and *Top2a* (cluster 5), and a cluster expressing high levels of ribosomal genes (Cluster 6). In addition, we obtained a cluster composed by both macrophages (∼64%) and monocytes (∼36%) characterized by the expression of *C1qa* and *C1qb* (cluster 1). Finally, among monocytes we detected classical monocytes expressing *Ccr2* and *Ly6c2* (cluster 3) and non-classical monocytes expressing *Ly6c2, Ace and Cd300e* (cluster 2), reproducing the immune landscape previously described in tumor-bearing mice(*69*). In monocytes/macrophages compartment from BAL patients, we observed 5 clusters (Fig. 4f): a macrophage cluster expressing proliferation markers *MKI67* and *TOP2A* (cluster d); a cluster characterized mainly by macrophages expressing *C1QA, C1QB* and *MRC1* (cluster a); a cluster expressing monocyte markers *FCN1* and *VCAN* (cluster b); two small clusters expressing ribosomial (cluster c) and heat shock protein (HSPs) genes (cluster e). Probably due to the intrinsic complexity(*69*) and the low number of cells in the mononuclear phagocyte compartment(*32*), we were not able to distinguish classical from non-classical monocytes as well as to identify different macrophage subsets in BAL samples. Therefore, unlike neutrophils, we could not reconstruct an effective map between mouse and human monocyte/macrophage subsets. However, we observed the expression of similar top marker genes between subset 5 and d, which identified proliferating macrophages (Supplementary information, Table S2); furthermore, the expression of several macrophage lineage genes such *C1QA, C1QB* and *MRC1* was shared between the two species. Conversely, inflammatory genes such as *IL1B, CXCL1* and *CXCL3* were mainly expressed by monocytes in BALs, unlike mouse dataset in which they were expressed at high levels both in monocyte and macrophages (Supplementary information, Table S2). Through gene set variation analysis (GSVA), we confirmed that inflammatory response and related pathways TNF-α signaling via NF-κB and JAK-STAT3 signaling were active in several clusters of mouse monocytes and macrophages while in BALs were specifically localized in monocytes (Fig. 4g). Despite several mouse subsets presented an up-regulation of inflammatory genes in vFLIP compared to WT mice (Fig. 4h, top panels) as well as high GSVA scores on inflammatory-related pathways (Fig. 4g), GSEA analysis at the cluster level unveiled significant up-regulation of inflammatory response, TNF-α signaling via NF-κB and JAK-STAT3 pathways specifically in cluster 4 (Supplementary information, Table S2), which was also the most dysregulated in terms of cell proportion (∼19% vFLIP; ∼8% WT) (Fig. 4e). Notably, the up-regulation of JAK-STAT3 signaling pathway, in vFLIP cluster 4 compared to WT, was paralleled by the expression of several genes in the pathway such as *Jak2, Csf1, Il10rb* and *Cd38* (Fig. 4h, bottom panels).

In summary, a conserved landscape of myeloid cells enriched for transcriptional signatures associated with the inflammatory response was detailed in the lung environment of both vFLIP mice and severe COVID-19 patients, in line with the concept that FLIP-expressing myeloid cells drive lung pathology.

### STAT3 targeting restrains immunopathology in vFLIP mice

To investigate whether targeting STAT3 could be sufficient to dampen the uncontrolled immune dysregulation in vFLIP mice, we evaluated therapeutic approaches involving either pharmacological or RNA interference of STAT3. Initially, we tested *in vivo* the efficacy of two STAT3 inhibitors: silibinin, a STAT3 inhibitor that blocks the Y705 phosphorylation-related and STAT3 dimerization(*70*), or baricitinib, a clinically-approved inhibitor of JAK1 and JAK2 able to interfere with STAT3 signaling activation(*71*). Four weeks after the establishment of BM chimerism in recipients (T0), mice were treated every two days by intraperitoneal injection. Weight loss was significantly decreased in mice that had received the drug treatments (Supplementary information, Fig. S5a). Furthermore, STAT3 targeting reduced the plasma concentration of several pro-inflammatory cytokines (Fig. 5a), which are produced at abnormal levels in untreated vFLIP mice and are also a feature of COVID-19(*3*). Immunohistochemistry of spleen demonstrated a reduction in systemic lymphopenia of treated vFLIP mice, with some differences, since baricitinib furthered a raise in both T and B cells while silibinin affected only T lymphocyte frequency (Fig. 5b). By analyzing eight different parameters (e.g. interstitial enlargement, vascular congestion, perivascular neutrophils, presence of thrombi, presence of infarction, fibroplasia, foam cell clusters and perivascular infiltrate; Supplementary information, Fig. S5b), we confirmed a reduction in the pathological score of inflammatory pneumonia in treated mice (Fig. 5c and Supplementary information, Fig. S5c).

**Fig. 5.**
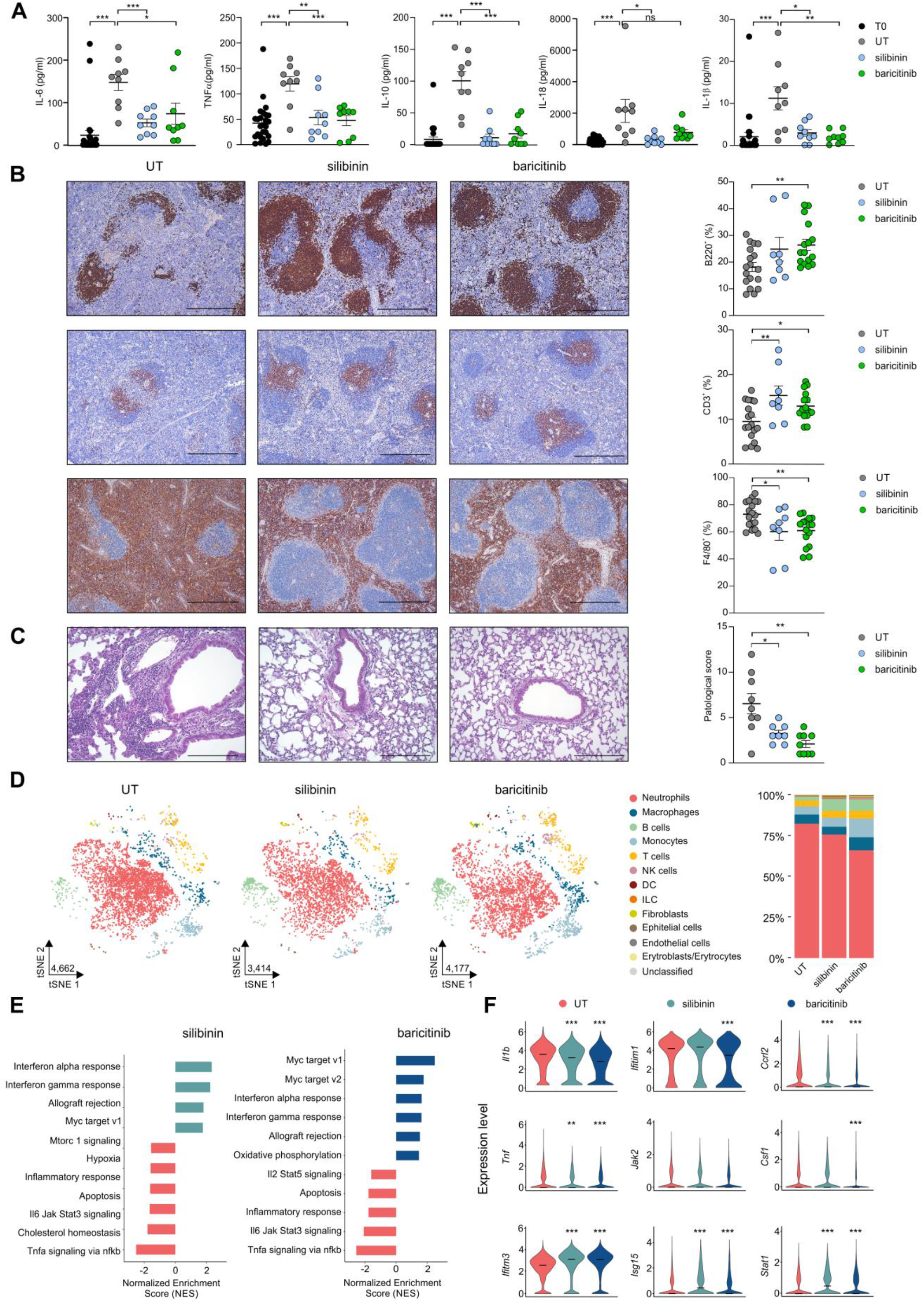
STAT3 pharmacological targeting reduces lung damage and immune dysfunctions in vFLIP mice. **(A)** Analysis of cytokines levels in serum of vFLIP mice before treatment (T0) or at the end of treatment (untreated, n=9; silibinin, n=9; baricitinib, n=9). **(B)** Lymphocytes and macrophages quantification in spleens of untreated (n=14), silibinin (n=8) and baricitinib (n=16) vFLIP mice by H&E staining. Scale bar, 200 μm. **(C)** Pathological score of lungs of untreated (n=14), silibinin (n=8) and baricitinib (n=16) vFLIP mice by H&E staining. Scale bar, 200 μm. **(D)** tSNE representation of scRNA-seq from untreated (4,662) mice and treated with silibinin (3,414) and baricitinib (4,177) colored according to cell type. Stacked bar plots representing cell type proportions across conditions. **(E)** Bar plot representing the up- and down-regulated (adjusted p-value < 0.05) hallmark gene sets obtained in the bulk-like analysis of treated compared to untreated vFLIP chimera cells obtained through GSEA analysis. **(F)** Violin plots showing the expression of genes involved in inflammatory response, JAK-STAT3 signaling pathway and interferon response in the lung of vFLIP chimera mice (*p < 0.05, **p < 0.01, ***p < 0.001). Data are reported as mean ± S.E.M. *p ≤ 0.05, **p ≤ 0.01 and ***p ≤ 0.001 by Student’s *t*-test, Mann–Whitney test.

To explore further the molecular underpinnings of the inflammatory shutdown, we evaluated the lung-infiltrating leukocyte profile of treated and untreated vFLIP mice by scRNA-seq. After projecting the cells into a 2-dimensional space using t-SNE, we evaluated the effect of pharmacological treatments on the proportions of the main cellular subsets. Both treatments reduced the neutrophil proportion (∼82% UT; ∼76% silibinin; ∼66% baricitinib), with a compensatory increase in other cells, such as lymphocytes, indicating a trend towards rebalancing the lung-infiltrating leukocyte frequency (Fig. 5d). Considering neutrophil subgroups, we observed a slight reduction in the major neutrophil subset (cluster 0: ∼89% UT; ∼87% silibinin; ∼86% baricitinib) in treated mice and an expansion in neutrophils expressing interferon-response genes (cluster 3: ∼2% UT; ∼4% silibinin: ∼5% baricitinib) (Supplementary information, Fig. S5d). Therefore, STAT3-targeting approaches can alter neutrophil composition favoring the accrual of elements with type I interferon response-associated genes. As for the monocytes/macrophages in treated mice, we observed a decrease in both classical monocytes (clusters 3: ∼32% UT; ∼23% silibinin; ∼25% baricitinib) and macrophages expressing inflammatory cytokines (cluster 4: ∼12% UT; ∼8% silibinin; ∼8% baricitinib); at the same time, we evinced the increase in clusters 1 (∼25% UT; ∼27% silibinin; ∼41% baricitinib) and cluster 2 (non-classical monocytes: ∼10% UT; ∼20% silibinin; ∼14% baricitinib) (Supplementary information, Fig. S5d).

Finally, GSEA analysis confirmed an up-regulation of interferon alpha/gamma pathways in both STAT3-based treatments compared to controls and, simultaneously, the down-regulation of gene signatures associated to inflammatory response, JAK-STAT3-dependent signaling pathways and TNF-α signaling via NF-κB (Fig. 5e). Consistent with this effect, several inflammatory genes, such as *Il1b, Clec5a, Ccrl2* and *Ifitm1* were down-regulated while *Ifitm3, Stat1* and *Isg15* genes, which are associated to interferon response pathways, were up-regulated in baricitinib-treated vFLIP mice (Fig. 5f). Thus, STAT3 inhibitors might mitigate the inflammatory pathology, both locally and systemically, by affecting the aberrant FLIPs-STAT3 feedforward loop while keeping the antiviral response active.

To confirm the immunomodulatory capacity of STAT3 inhibitors, we tested the ability of these two compounds to control the immunosuppressive functions of c-FLIP-expressing monocytes isolated from COVID-19 patients. T cell proliferation was significantly preserved after co-culture with monocytes pre-treated with the two drugs, as compared to untreated controls (Supplementary information, Fig. S5e). These results indicate that STAT3-targeting may prevent T cell dysregulations by limiting immunosuppressive features of SARS-CoV-2-educated myeloid cells, endorsing the clinical results about baricitinib efficacy in altering immunoregulatory properties of myeloid cells in COVID-19 patients(*59*).

To provide complementary evidence that a direct STAT3-silencing in myeloid cells could control the evolution of immunopathological disorders in vFLIP mice, we exploited the *in vivo* delivery of 4PD nanoparticles loaded with STAT3-specific short hairpin RNAs (shSTAT3) on their surface. The ability of 4PDs to recognize and target preferentially mononuclear phagocytes and mediate an effective *in vivo* delivery of shSTAT3 was previously proven in cancer mouse models(*72*). Therefore, vFLIP mice were treated by a total of nine administrations, every two days, of 4PDs loaded with either a scrambled RNA sequence (shCTRL, as negative control) or shSTAT3. Similar to STAT3 pharmacological treatment, the genetically-based STAT3 silencing in myeloid compartment controlled the weight loss, limiting CRS-associated cachexia (Supplementary information, Fig. S6a). Moreover, STAT3-silencing affected the severity of inflammatory pneumonia, as highlighted by the decrease in lung pathological score (Fig. 6a and Supplementary information, Fig. S6b) and allowed a complete contraction of neutrophilia in pulmonary tissues (Fig. 6b and Supplementary information, Fig. S6c). Notably, shSTAT3 treated mice showed also a reduction in systemic lymphopenia, with an increased frequency of both T and B lymphocytes and a concomitant myeloid cell reduction in the spleen (Fig. 6c) and in peripheral blood (Fig. 6d). Finally, the 4PD-mediated, STAT3-specific shRNA delivery normalized the plasma concentration of several pro-inflammatory cytokines (Fig. 6e).

**Fig. 6.**
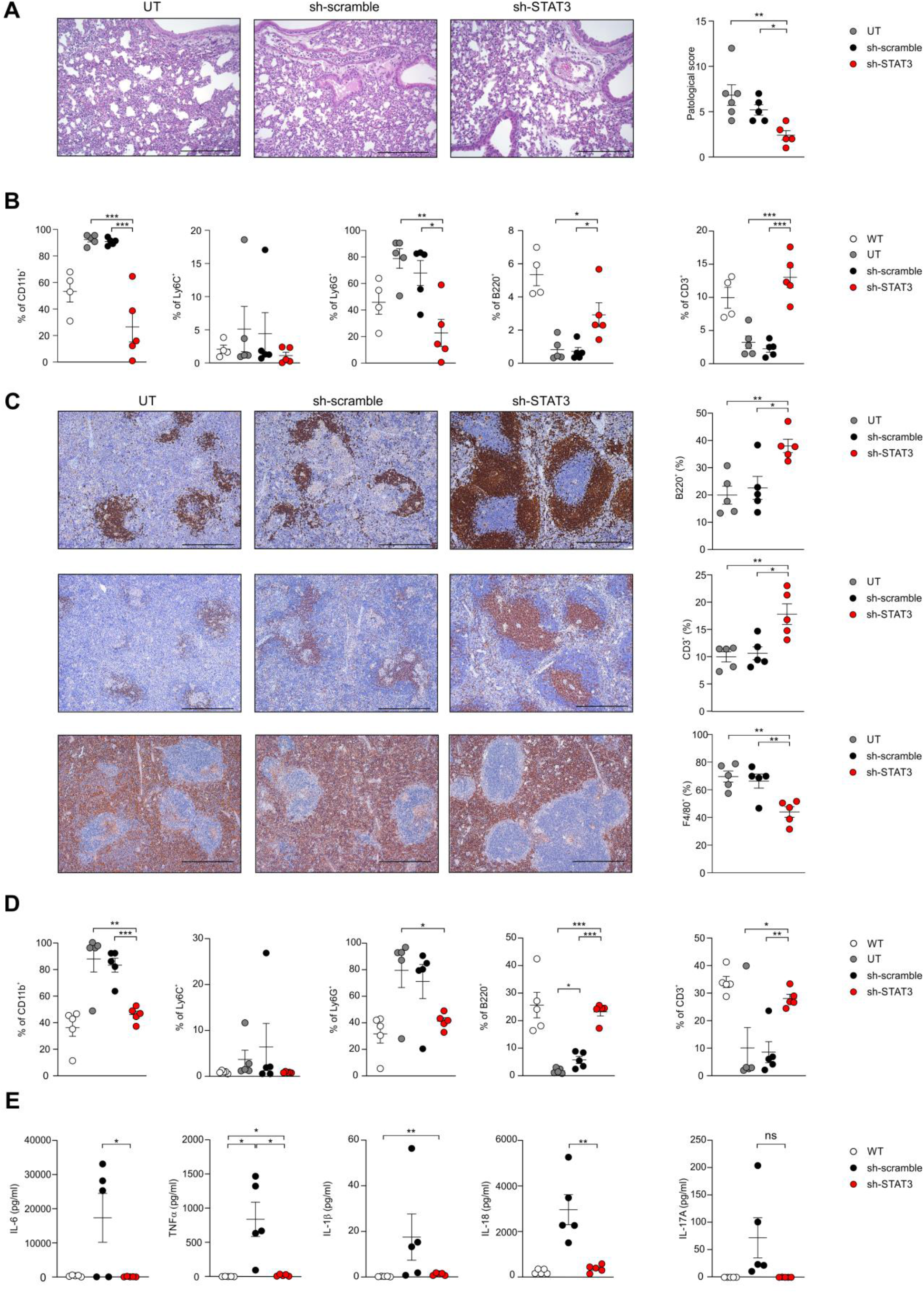
*In vivo* STAT3-silencing approach mitigates the evolution of immunopathological disorders in vFLIP mice. **(A)** Pathological score of lungs of vFLIP mice (untreated, n=6; sh-scramble, n=5; sh-STAT3, n=5) by H&E staining. Scale bar, 200 μm. **(B)** Flow cytometry analysis of CD45^+^ leukocytes isolated from lungs of vFLIP mice (untreated, n=6; sh-scramble, n=5; sh-STAT3, n=5) or WT mice (n=5). **(C)** Lymphocytes and macrophages quantification in spleens of vFLIP mice (untreated, n=6; sh-scramble, n=5; sh-STAT3, n=5) by H&E staining. Scale bar, 400 μm. **(D)** Flow cytometry analysis in peripheral blood in vFLIP mice (untreated, n=6; sh-scramble, n=5; sh-STAT3, n=5) or WT mice (n=5). **e** Analysis of cytokines levels in serum of WT (n=5), sh-scramble (n=5) or sh-STAT3 (n=5) vFLIP mice. Data are reported as mean ± S.E.M. *p ≤ 0.05, **p ≤ 0.01 and ***p ≤ 0.001 by Mann– Whitney test.

Taken together, these data indicate that STAT3-targeting, especially in myeloid cells, is effective in tempering CRS-associated immunopathological disorders triggered by the accumulation of FLIP-expressing cells.

## Discussion

To date, the pathology of CRS is incompletely understood and no single definition is widely accepted by the scientific community. Our data establish FLIP-expressing myeloid cells as a pivotal driver of CRS. Indeed, the vFLIP chimera mice show acute systemic inflammatory symptoms (Fig. 2 and Supplementary information, Fig. S2-3), elevated cytokine levels (Fig. 5a), weight loss (Supplementary information, Fig. S5a), lymphopenia (Supplementary information, Fig. S2c), lung injury (Fig. 2a) endothelial cell activation (Fig. 2d) and multiple organ dysfunctions (Supplementary information, Fig. S2d). The impact of vFLIP activation on building up inflammatory diseases is confirmed also by evidence about the role of this anti-apoptotic protein in inflammatory bowel disease(*73*), multicentric Castleman disease in mice(*74*), as well as on reprogramming myeloid cells into immunosuppressive elements in cancer(*35*).

A significant gap remains between pre‐clinical testing and clinical trials to treat efficiently CRS, as well as to identify key molecular mechanisms that control the pathogenic process. In the last decades, mouse models of sepsis, such as LPS-induced inflammation, or the development of humanized mice based on the engraftment of human PBMCs into immunodeficient mice, which mimic CAR T-cell therapy-induced cytokine storm, have been primarily developed to replicate clinical stages and outcomes of CRS, as platforms for screening potential therapies(*6, 75*). However, no suitable and appropriate experimental *in vivo* models have been developed to identify alterations in molecular and cellular processes that might highlight the triggers of CRS. Our study based on a tissue-specific transgenic, conditional knock-in mouse model offer a novel tool for defining the mechanisms that fuel inflammation and CRS-associated immune dysfunctions.

In line with our results, recent studies underlined that c-FLIP expression was enhanced in several SARS-CoV-2 infected cell lines, where the expression of FLIP suppressors, such as the forkhead transcriptional factor FoxO3A(*76*), was abrogated(*30, 77*). Since the interaction of c-FLIP to FADD and/or caspase-8 or -10 and TRAIL receptor 5 prevents death-inducing signaling complex formation and subsequent activation of the caspase cascade(*78*), it is plausible that SARS-CoV-2 virus exploits FLIP-mediated cell death delay for its own replication. Our analysis of both SARS-CoV-2-infected hACE2 transgenic mice and autopsy samples from lung in COVID-19 patients demonstrated, for the first time, the *in vivo* overexpression of FLIP in myeloid cells in severe COVID-19 (Fig. 1). However, compared to the traditionally ascribed FLIP intervention on cell-survival, our findings rather point to the additional function of modulating myeloid cells to determine CRS progression (Fig. 1d-e; Fig. 2i).

Monocytes from both COVID-19 patients (Fig. 1c) and vFLIP transgenic mice(*35*) display immunosuppressive properties and are a source of pro-inflammatory cytokines (Fig. 1e and Supplementary information, Fig. S1d). We found that immunoregulatory functions of monocytes isolated from COVID-19 patients correlated with the expression of PD-L1 (Supplementary information, Fig. S1c), suggesting a possible contribution of immune checkpoint engagement on T cell blockade during COVID-19 evolution. These data are in line with our previous findings showing that FLIP-expressing monocytes isolated from PDAC patients had high levels of surface PD-L1(*35*) and confirm previous reports indicating that immunosuppressive myeloid cells in COVID-19 patients did exhibit increased *CD274* mRNA levels(*32, 34*). In agreement with Schulte-Schrepping’s report in which STAT3 was suspected as transcriptional factor of immunosuppressive monocytes in COVID-19, we demonstrated that immunosuppression by monocytes isolated from COVID-19 patients can be indeed reverted by STAT3 inhibitors (Supplementary information, Fig. S5e). Furthermore, we unveiled a concomitant expression of FLIP and activated STAT3 signaling in myeloid cells of SARS-CoV-2 infected hosts (Fig. 1a and Fig. 1f), as well as in vFLIP transgenic mouse model (Fig. 2c). Further investigations should mechanistically address the unconventional properties of c-FLIP as transcription factor, either by itself or cofactor of transcriptional machinery capable of activating STAT3 signaling pathway in myeloid cells under pathological conditions, which to date are only partially demonstrated in immortalized cell lines(*43, 79*).

Since the *in vitro* enforced expression of FLIP in myeloid cells promotes the overexpression of several pro-inflammatory cytokines (i.e. IL-6, IL-7, IL-10, CSF3 and TNF-α) by a “steered” NF-κB activation, which also results in enhanced STAT3-signalling activation(*35*), we argue that a pervasive inflammatory loop is established by FLIP through the joint action of NF-κB and STAT3 during CRS evolution. Indeed, a synergy between NF-κB and STAT3 molecules based on pro-inflammatory cytokines (i.e. IL-6), which act as inflammation amplifier, was reported in several multiple inflammatory and autoimmune diseases(*80*) and postulated also in COVID-19(*81*). In agreement with this paradigm, a recent analysis of multi-organ proteomic landscape of COVID-19 autopsies confirmed both NF-κB and STAT3 as transcription factors largely upregulated in multiple organs (*82*), implying a widespread and pervasive activations of the two pathways.

Our *in vivo* findings show that STAT3-targeting provides a significant disease control in mice with CRS, unexpectedly highlighting how blocking a single member of the NF-κB/STAT3 loop is sufficient to halt pathological inflammation. Data presented here are in line with recent clinical results about baricitinib efficacy in controlling SARS-CoV-2-mediated immune dysregulation(*59, 68, 83, 84*) and with the decision of U.S. Food and Drug Administration to approve baricitinib in combination with remdesivir for the treatment of severe COVID-19 patients. More important, we demonstrated that Jak1/Jak2 inhibitor did not affect negatively genes associated with type I IFN antiviral responses but, on the contrary, pharmacological treatment was associated with a relative increase in interferon-stimulated genes (Fig. 5e-f), likely by re-programming the myeloid composition in the lung environment (Fig. 5d and Supplementary information, Fig. S5c). This signaling switch might be due to the activity of STAT1 occupying space on STAT3-activating receptors, as suggested by the conversion of IL-6R signaling to a dominant STAT1 activation in STAT3-deficient cells(*85, 86*). Finally, the striking results on the normalization of the immune landscape, organ pathology and cachexia following shSTAT3-based treatment (Fig. 6 and Supplementary information, Fig. S6) clearly finger at STAT3 as the main target in FLIP expressing myeloid cells and define it as the most deleterious cause of immune and tissue damage during CRS.

Despite the caveats linked to species-specific profiles, our findings revealed some conserved genetic features of lung-infiltrating myeloid cells between vFLIP mice and COVID-19 patients. Indeed, neutrophil subsets characterized by the expression of S100-family genes, type I interferon response genes (ISG15/ISG20) and chemokines (*Ccl3/CCL3*), as well as macrophages expressing proliferation-associated gene signatures (*Mki67, Top2a*) showed similarities among species. The shared leukocyte subsets showed a higher expression of inflammatory response-associated genes and were more prone to STAT3 therapy.

In summary, in this study we demonstrated the pivotal role of FLIP-expressing myeloid cells to stimulate directly a lethal inflammatory status, by fueling an aberrant STAT3-dependent signaling pathway. Moreover, we substantiated the therapeutic effectiveness of STAT3 on-target strategy to mitigate uncontrolled inflammation and acute disease, which serve as a foundation for the development of more accurate and evidence-based therapies to control CRS disorders, as well as severe clinical aspects of the ongoing COVID-19 pandemic crisis.

## Materials and Methods

### Study design for patients

This study was designed to explore the impact of FLIP/STAT3 axis in promoting immunological alteration in COVID-19 patients. All 48 patients with COVID-19 and 4 healthy donors in this study were admitted, within the period from March 12th to April 20th 2020 to the University Hospital of Verona or Hospital of Pescara. At sampling, the stage of disease was categorized as mild (patients not requiring non-invasive/mechanical ventilation and/or admission to ICU) or severe (patients requiring admission to ICU and/or non-invasive/mechanical ventilation).

For immunohistochemistry analysis of lung autopsy, this study includes a group of 4 non-respiratory disease (NDR) patients, 4 bacterial pneumonia (BD) patients and 23 COVID-19 patients. The clinical features are recapitulated in Supplementary information, Table S1a-b.

For molecular data (i.e. single cell transcriptomic analysis), phenotypic analysis (myeloid characterization in terms of expression of immune suppression hallmarks) and functional data (myeloid immune suppressive assay), this study includes a group of 14 severe COVID-19 patients admitted to ICU, 11 mild SARS-CoV-2 patients and 4 HDs. The clinical features of the 3 groups of subjects are recapitulated in Supplementary information, Table S1c.

All the patients (and/or initially their families) provided written informed consent before sampling and for the use of their clinical and biological data. This study was approved by the local ethical committee (protocol 17963; principal investigator, Vincenzo Bronte; ClinicalTrials.gov identifier NCT04438629). All clinical investigations were conducted according to Declaration of Helsinki principles, and informed consent was obtained from all study participants.

### Study design for mice

This study was designed to define vFLIP^+^p-STAT3^+^myeloid cells as a main driver for CRS progression as well as to test STAT3-targeting approach as a possible strategy to ameliorate CRS-undergoing hosts. All genetically transgenic mice and their respective controls were gender and age-matched (typically 3–10 weeks) and both males and females were used in this study. Mice were assigned randomly to experimental groups. Germ free C57BL/6 mice were originally purchased from Charles River Laboratories Inc., CD45.1^+^congenic mice (*B6*.*SJL-Ptrc*^*a*^*Pepc*^*b*^*/BoyJ*) and LySM-CRE mice (*B6*.*129P2-Lyz2*^*tm1(cre)Ifo*^*/J*) were purchased from Jackson Laboratories; Rosa26.vFLIP mice were a gift from Dr. Ethel Cesarman (Weill Cornell Medicine, NY, USA).

To generate the vFLIP-chimera mouse model (hereafter named vFLIP mice), C57BL/6 female of 8 weeks of age received 9 Gy total body irradiation (TBI) using ^137^Cs-source irradiator. Six hours after pre-conditioning, irradiated recipient mice were intravenously injected with 5×10^6^ bone marrow cells obtained from CD45.1 WT and ROSAvFLIP Tg (CD45.2) donor mice at different ratio. Bone marrow cells over-expressing FLIP protein from Kaposi’s sarcoma virus (v-FLIP) in myeloid compartment were collected from ROSA26.vFLIP Tg knock-in mice. These mice were obtained by crossing ROSA26.vFLIP knock-in mice with mice expressing Cre recombinase under control of the endogenous *Lyz2* promoter. For the therapeutic studies, 50% WT-50% vFLIP ratio was used to generate the vFLIP-chimera mice. Four weeks post bone-marrow transplantation, peripheral blood of recipient mice was analyzed for the presence of donor-derived cells.

The *in vivo* effect of drugs treatment was investigated in the vFLIP-chimera mouse model, four weeks after the bone marrow cells transplantation. Chimera mice that displayed at least 20% of donor-derived cells were randomized before beginning treatment. Chimera mice were treated using 8 intraperitoneal administrations of Baricitinib (10 mg/kg; Cayman chemicals) or Silibinin (100mg/kg; Sigma-Aldrich) every two days, for a total of 9 treatments. shRNA treatments were performed three times a week by injecting shRNA (anti-STAT3 or scramble) loaded onto 4PD intravenously as previously described(*72*), for a total of 9 treatments. shRNA STAT3 sequence: 5’-UAAUACGACUCACUAUAAGGAGGGUGUCAGAUCACAUGGGCUUUCAAGAGAUCUCAACGGA C AUGCUACUGCCUU-3’; shRNA scramble STAT3 sequence: 5’-UAAUACGACUCACUAUAAGGGCAGUAGCAUGGUCCGUUGAGAUUCAAGAGAUCUCAACGGA C AUGCUACUGCCUU-3’ (Boston Open Labs). Immunohistochemistry (IHC) and flow cytometry analysis were performed at the end of the experiment (2 weeks after the first treatment). Chimera mice were euthanized when the weight loss reached the 20% of body-weight as an animal protocol-defined endpoint.

All mice were maintained under specific pathogen-free conditions in the animal facility of the University of Verona. Food and water were provided *ad libitum*. Animal experiments were performed according to national (protocol number C46F4.26 approved by the Ministerial Decree Number 993/2020-PR of July 24, 2020 (PI: Stefano Ugel) and protocol number C46F4.8 approved by the Ministerial Decree Number 207/2018-PR of February 21, 2018) and European laws and regulations. All animal experiments were approved by Verona University Ethical Committee and conducted according to the guidelines of Federation of European Laboratory Animal Science Association (FELASA). All animal experiments were in accordance with the Amsterdam Protocol on animal protection and welfare: mice were monitored daily and euthanized when displaying excessive discomfort.

### Statistical analysis

All data are reported as mean ± standard error (SE) of the mean. Statistical analyses were performed using Graph Pad Prism (version 8.0.2). Student *t* test (parametric groups) and Wilcoxon–Mann–Whitney test (nonparametric groups) were used to determine statistical significance of differences between two treatment groups. Values were considered significant at *P* ≤ 0.05.

## Data Availability

ScRNA-seq data generated in this study are deposited in the Gene Expression Omnibus (GEO) under the accession number GSE168098.

## Acknowledgments

We thank Silvia Sartoris, Rosalinda Trovato, Varvara Petrova, Laura Pinton, Monica Castellucci, Tommaso Grandi and Gianmarco Tarquini for their support and helpful discussion during the development of this research. We thank Giulio Fracasso, Ornella Poffe, Cristina Anselmi and Tiziana Cestari for perfect technical assistance. We thank Pierre Bost, David Eyal, Jimmy Caroli, Oriana Romano and Mattia Forcato for bioinformatics support. We deeply acknowledge the contribution of ‘Centro Piattaforme Tecnologiche’ of the University of Verona for sorting and imaging experiments and ‘Centro Interdipartimentale di Servizio alla Ricerca Sperimentale’ of the University of Verona for maintaining mouse colonies. We thank Ethel Cesarman (Weill Cornell Medical College, New York, NY, USA) for the gift of the Rosa26.vFLIP conditional allele mice. We also thank all the personnel involved in the patients’ care and assistance; in particular, we thank Sara Boschetti, Francesca Del Favero, Riccardo Boetti, Pasquale De Nardo, Fabiana Busti, Chiara Mozzin, Federica Marola and Valentina Albanese. We wish to thank all the members of the Immunology section of Department of Medicine of University of Verona who worked forcefully during pandemic. We dedicate this work to the memory of the health care workers who have given their lives in the care of patients with COVID-19.

## Author Contributions

Conceptualization: C.M., Si.C., A.F., V.B., S.U.;

Methodology: C.M., Si.C., A.F. and S.U.;

Investigation: C.M., Si.C., A.F., C.F., F.D.S., St.C., A.A., F.H., R.M.B. and S.U.;

Immunohistochemistry and pathology analysis, data interpretation: A.L., F.D.P. and M.I.; Computational analysis and data interpretation: Si.C., A.G. and S.B.;

Methodology for the in vivo RNA delivery experiments and data interpretation: S.Z. and P.S.;

Responsible for biological specimens and patients’ data collection: E.T., K.D., L.G., E.P., D.G., I.P., P.A.I., D.A. and A.C;

Methodology of human ACE2 mouse model, pathological analysis and data interpretation: Y.C. and Z.L.S.

Writing and data interpretation: P.J.M, M.C. and I.A., V.B and S.U.;

Supervision: V.B. and S.U.;

Funding acquisition: V.B. and S.U.

