## Supplementary material for "Fatal cytokine release syndrome by an aberrant FLIP/STAT3 axis": Supplmentary file

**SUPPLEMENTAL INFORMATION**

**Supplemental Figures**

- Fig. S1. Clinical characteristic of enrolled patients and immunosuppressive function of monocytes.
- Fig. S2. vFLIP chimera mice develop cytokine release syndrome.
- Fig. S3. Systemic and local features of cytokine release syndrome in vFLIP mice.
- Fig. S4. Single-cell transcriptional profiling of lung-infiltrating cells in vFLIP mice and BAL-derived immune cells obtained from COVID-19 patients.
- Fig. S5. Characterization of the pharmacological STAT3-targeting effectiveness in vFLIP mice.
- Fig. S6. Characterization of the in vivo STAT3-silencing effectiveness in vFLIP mice.
- Table S1. Patients characteristics.
- Table S2. scRNA-seq data and statistics.

**A**

**Main histopatological data of the study population.**

| Parameters | Total (N=23) |
|---|---|
| **Airways** | |
| Tracheobronchial inflammation, n (%) | |
| Acute, diffuse | 1 (4.3%) |
| Acute, focal | 1 (4.3%) |
| Chronic, diffuse | 0 |
| Chronic, focal | 1 (4.3%) |
| Not avaiable | 20 (87.1%) |
| **Alvelolar space** | |
| Acute inflammation, n (%) | |
| Present, diffuse | 1 (4.3%) |
| Present, focal | 5 (21.7%) |
| Absent | 17 (74%) |
| Chronic inflammation, n (%) | |
| Present, diffuse | 1 (4.3%) |
| Present, focal | 8 (34.8%) |
| Absent | 14 (60.9%) |
| Hyaline membrane, n (%) | |
| Present, diffuse | 6 (26.1%) |
| Present, focal | 1 (4.3%) |
| Absent | 16 (69.6%) |
| Pneumocyte type II hyperplasia, n (%) | |
| Present, diffuse | 2 (8.7%) |
| Present, focal | 1 (4.3%) |
| Absent | 20 (87%) |

| Parameters | Total (N=23) |
|---|---|
| **Alveolar wall** | |
| Chronic inflammation, n (%) | |
| Present, diffuse | 4 (17.4%) |
| Present, focal | 15 (62.2%) |
| Absent | 4 (17.4%) |
| **Fibrosis, n (%)** | |
| Present, focal | 11 (47.8%) |
| Present, diffuse | 9 (39.1%) |
| Absent | 3 (13.1%) |
| **Vessels, n (%)** | |
| Vessel congestion | 7 (30.4%) |
| Blood extravasation | 6 (26.1%) |
| Microthrombi | 1 (4.3%) |
| Large thrombi | 3 (13%) |
| **Other lesions, n (%)** | |
| Succular emphysema | 6 (26.1%) |
| Calcifications | 2 (8.7%) |
| Anthracosis | 3 (13%) |
| Myofibroblastic foci | 2 (8.7%) |

**B**

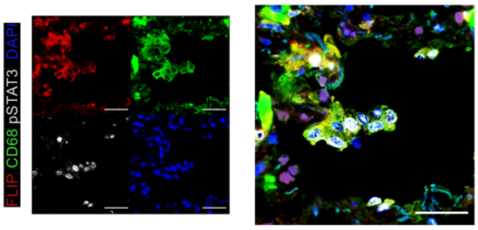

**C**

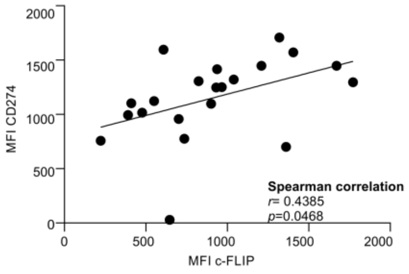

**D**

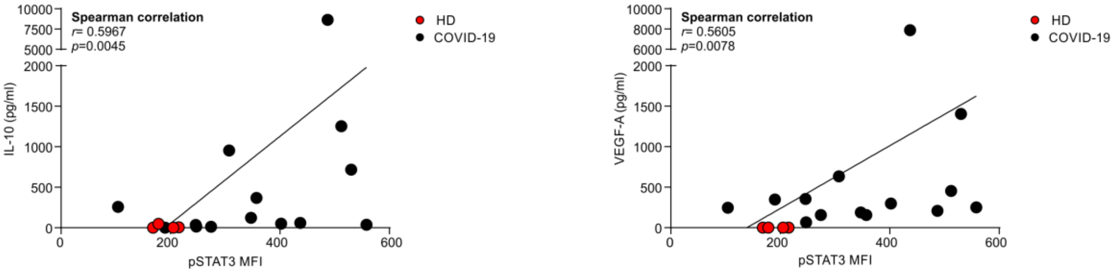

**Fig. S1. Clinical characteristic of enrolled patients and immunosuppressive function of monocytes. (A)** Lung histopathological characteristics of COVID-19 patients. **(B)** Representative immunofluorescence (IF) staining of monocytes of COVID-19 patient. Cells were stained for CD68 (green), c-FLIP (red), pSTAT3 (white) and DAPI (blue). Scale bar, 20 μm. **(C)** Correlation between CD274 and c-FLIP expression in COVID-19 $CD14^{+}$ cells (n=19). **(D)** Correlation between the release of IL-10 or VEGF-A cytokines and pSTAT3 expression in circulating $CD14^{+}$ cells from HD (red, n=4) and COVID-19 patients (black, n=13). Correlation analysis was performed by Spearman's rank correlation (**C**, **D**).

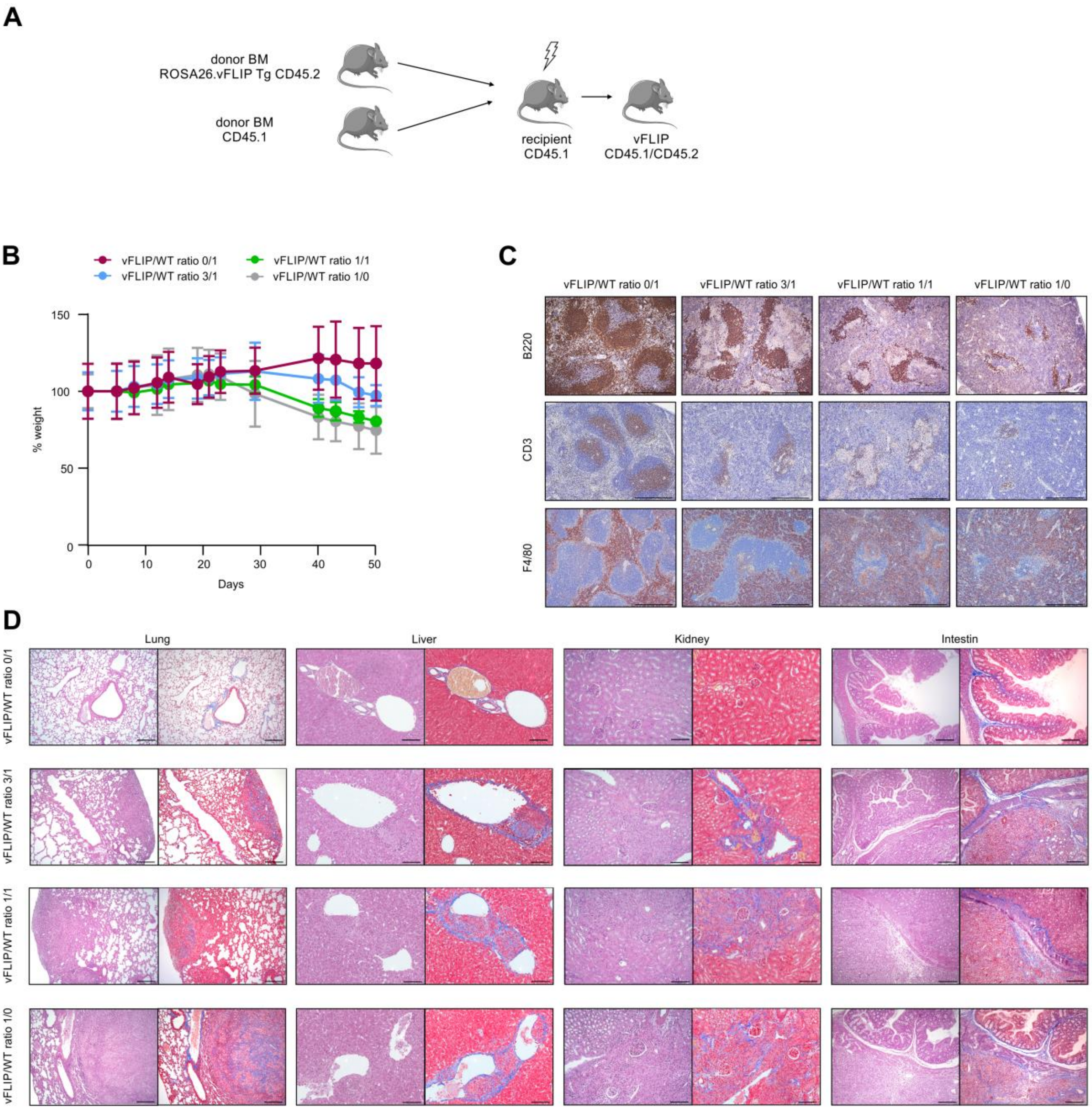
A
donor BM
ROSA26.vFLIP Tg CD45.2
donor BM
CD45.1
recipient
CD45.1
vFLIP
CD45.1/CD45.2
B
vFLIP/WT ratio 0/1
vFLIP/WT ratio 3/1
vFLIP/WT ratio 1/1
vFLIP/WT ratio 1/0
% weight
150
100
50
0
0
10
20
30
40
50
Days
C
vFLIP/WT ratio 0/1
vFLIP/WT ratio 3/1
vFLIP/WT ratio 1/1
vFLIP/WT ratio 1/0
B220
CD3
F4/80
D
Lung
Liver
Kidney
Intestin
vFLIP/WT ratio 0/1
vFLIP/WT ratio 3/1
vFLIP/WT ratio 1/1
vFLIP/WT ratio 1/0

**Fig. S2. vFLIP chimera mice develop cytokine release syndrome.**

**(A)** Experimental layout of chimera (vFLIP) mice generation. **(B)** Variation of body weight during CRS progression in vFLIP chimera mice (n=3, for each condition). **(C)** IHC analysis of spleen in vFLIP mice. Scale bar, 400 µm. **(D)** Representative H&E-stained microscopy images and Masson's Trichrome of lung, liver, kidney and intestine of vFLIP mice. Scale bar, 100 µm.

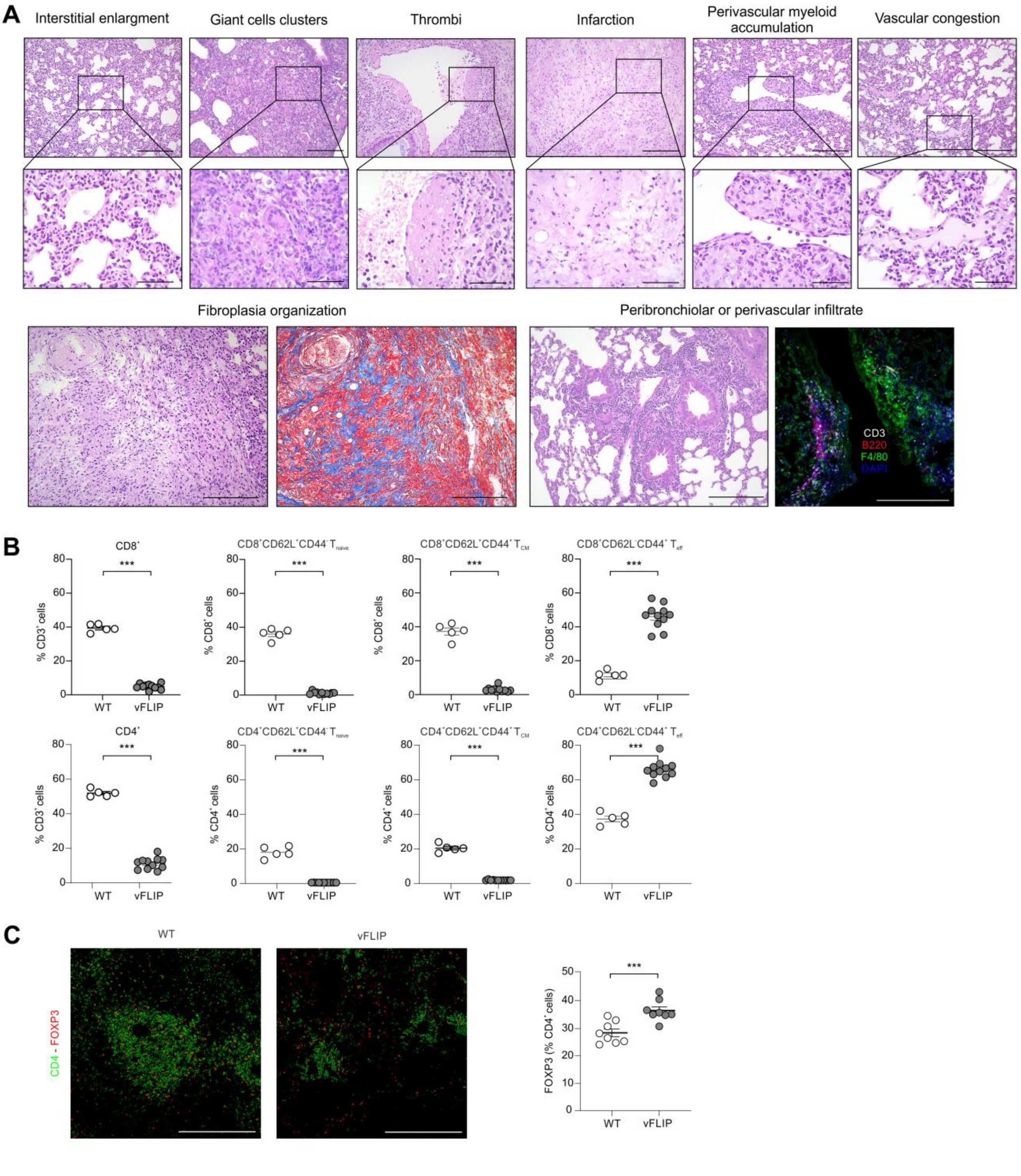
A
Interstitial enlargment
Giant cells clusters
Thrombi
Infarction
Perivascular myeloid accumulation
Vascular congestion
Fibroplasia organization
Peribronchiolar or perivascular infiltrate
CD3
B220
F4/80
DAPI
B
$CD8^+$
$CD8^+CD62L^+CD44^-\ T_{naive}$
$CD8^+CD62L^+CD44^+\ T_{CM}$
$CD8^+CD62L^-CD44^+\ T_{eff}$
$CD4^+$
$CD4^+CD62L^+CD44^-\ T_{naive}$
$CD4^+CD62L^+CD44^+\ T_{CM}$
$CD4^+CD62L^-CD44^+\ T_{eff}$
% $CD3^+$ cells
% $CD8^+$ cells
% $CD4^+$ cells
***
0
20
40
60
80
WT
vFLIP
C
WT
vFLIP
CD4 - FOXP3
FOXP3 (% $CD4^+$ cells)
0
10
20
30
40
50
***
WT
vFLIP

**Fig. S3. Systemic and local features of cytokine release syndrome in vFLIP mice.**

**(A)** Representative H&E-stained microscopy images of pathological score: interstitial enlargement, giant cells clusters, thrombi, infarction, perivascular myeloid accumulation, vascular congestion, fibroplasia organization and peribronchiolar or perivascular infiltrate vFLIP mice. Scale bar, 200 µm (upper panel) and 50 µm (bottom panel). **(B)** Flow cytometry analysis of $CD3^+$ T cell subsets in the spleen of vFLIP mice (n=11) and WT mice (n=5). **(C)** Representative IF staining of $CD4^+$ $FOXP3^+$ cells in spleen of vFLIP mice (n=8) and WT (n=8). Data are reported as mean ± S.E.M. *$p \leq 0.05$, **$p \leq 0.01$ and ***$p \leq 0.001$ by Mann–Whitney test (**B**,**C**).

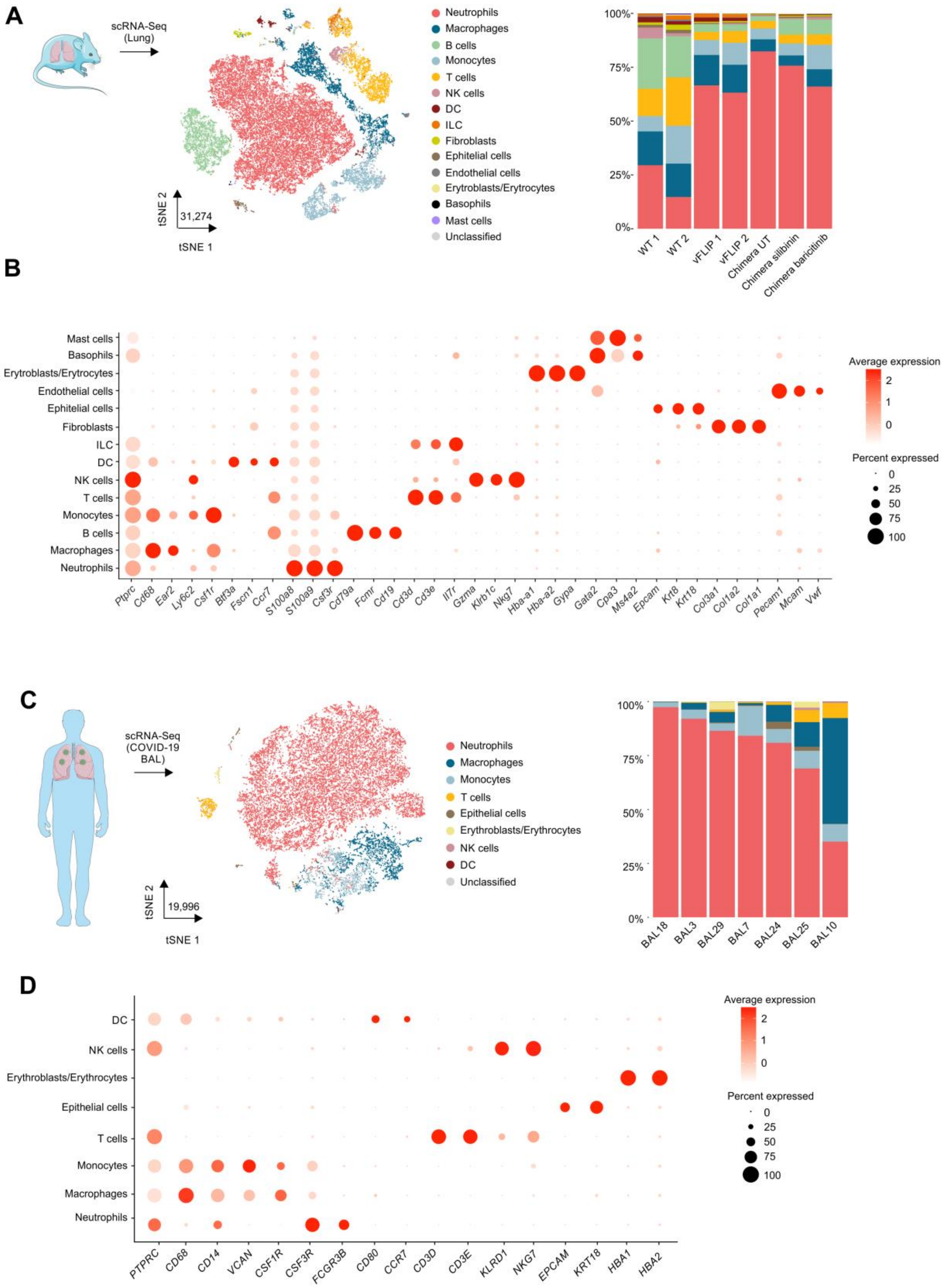
A
scRNA-Seq (Lung)
tSNE 2
31,274
tSNE 1
Neutrophils
Macrophages
B cells
Monocytes
T cells
NK cells
DC
ILC
Fibroblasts
Ephitelial cells
Endothelial cells
Erytroblasts/Erytrocytes
Basophils
Mast cells
Unclassified
100%
75%
50%
25%
0%
WT 1
WT 2
vFLIP 1
vFLIP 2
Chimera UT
Chimera silibinin
Chimera baricitinib
B
Mast cells
Basophils
Erytroblasts/Erytrocytes
Endothelial cells
Ephitelial cells
Fibroblasts
ILC
DC
NK cells
T cells
Monocytes
B cells
Macrophages
Neutrophils
Ptprc
Cd68
Ear2
Ly6c2
Csf1r
Batf3a
Fscn1
Ccr7
S100a8
S100a9
Csf3r
Cd79a
Fcmr
Cd19
Cd3d
Cd3e
Il7r
Gzma
Klrb1c
Nkg7
Hba-a1
Hba-a2
Gypa
Gata2
Cpa3
Ms4a2
Epcam
Krt8
Krt18
Col3a1
Col1a2
Col1a1
Pecam1
Mcam
Vwf
Average expression
2
1
0
Percent expressed
0
25
50
75
100
C
scRNA-Seq (COVID-19 BAL)
tSNE 2
19,996
tSNE 1
Neutrophils
Macrophages
Monocytes
T cells
Epithelial cells
Erythroblasts/Erythrocytes
NK cells
DC
Unclassified
100%
75%
50%
25%
0%
BAL18
BAL3
BAL29
BAL7
BAL24
BAL25
BAL10
D
DC
NK cells
Erythroblasts/Erythrocytes
Epithelial cells
T cells
Monocytes
Macrophages
Neutrophils
PTPRC
CD68
CD14
VCAN
CSF1R
CSF3R
FCGR3B
CD80
CCR7
CD3D
CD3E
KLRD1
NKG7
EPCAM
KRT18
HBA1
HBA2
Average expression
2
1
0
Percent expressed
0
25
50
75
100

**Fig. S4. Single-cell transcriptional profiling of lung-infiltrating cells in vFLIP mice and BAL-derived immune cells obtained from COVID-19 patients.**

**(A)** tSNE representation of scRNA-seq from all mouse samples (31,274 cells) colored according to cell type. Stacked bar plots representing cell type proportions across all the mouse samples. **(B)** Dot plot showing the scaled average expression of known marker genes for the mouse cell populations identified. **(C)** tSNE representation of scRNA-seq from fatal COVID-19 BALs patients (19,996) colored according to cell type. Stacked bar plots representing cell type proportions across all the human BAL samples. **(D)** Dot plot showing the scaled average expression of known marker genes for the cell types identified in the BALs.

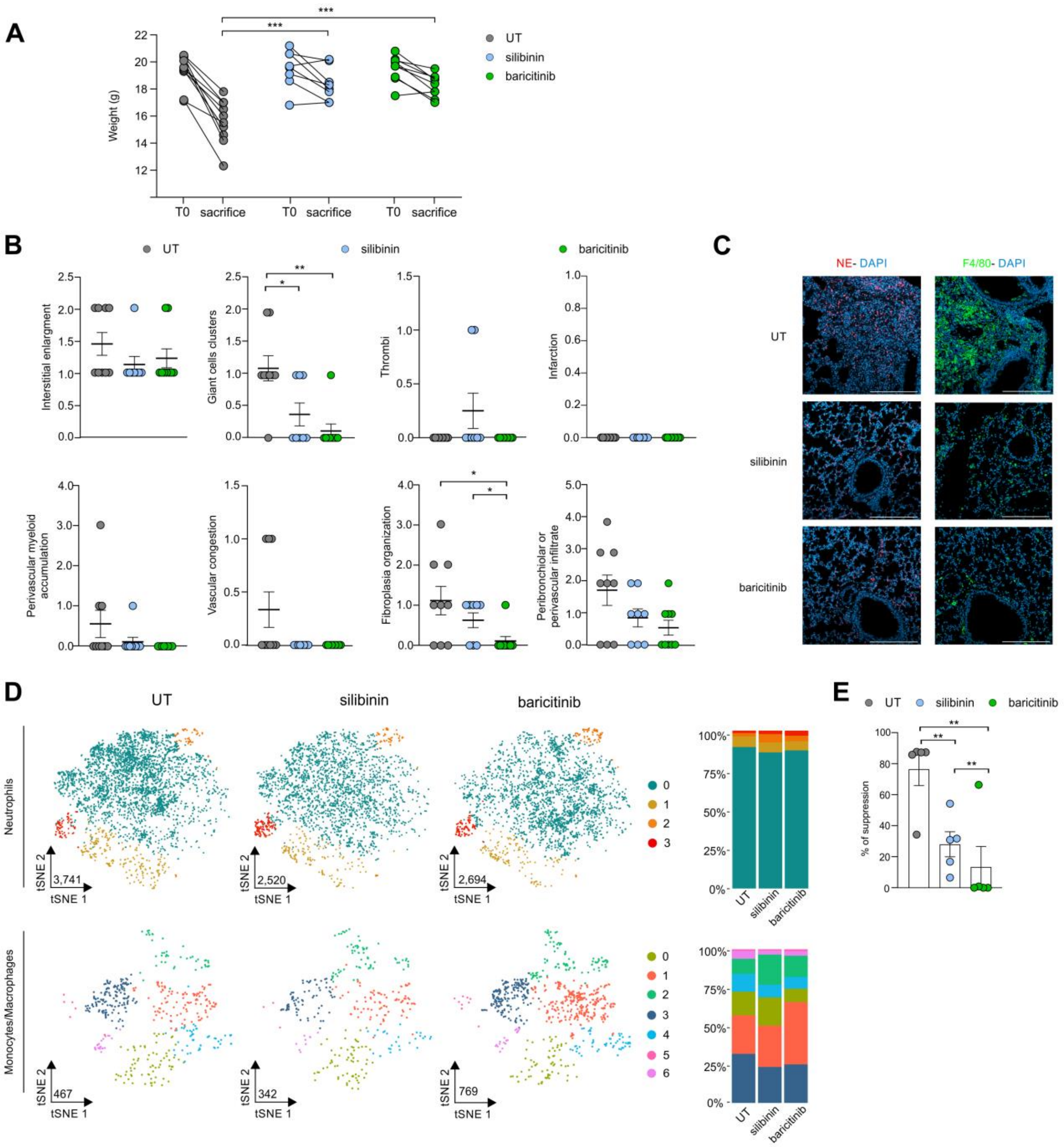
A
Weight (g)
T0
sacrifice
***
UT
silibinin
baricitinib
B
Interstitial enlargment
Giant cells clusters
Thrombi
Infarction
Perivascular myeloid accumulation
Vascular congestion
Fibroplasia organization
Peribronchiolar or perivascular infiltrate
C
NE- DAPI
F4/80- DAPI
D
Neutrophils
Monocytes/Macrophages
tSNE 1
tSNE 2
3,741
2,520
2,694
467
342
769
E
% of suppression
**

**Fig. S5. Characterization of the pharmacological STAT3-targeting effectiveness in vFLIP mice.**

**(A)** Body weights of vFLIP chimera mice before treatment (T0) or at the end of treatment (untreated, n=9; silibinin, n=8; baricitinib, n=9). **(B)** Dot plots of pathological score parameters: interstitial enlargement, giant cells clusters, thrombi, infarction, perivascular myeloid accumulation, vascular congestion, fibroplasia organization and peribronchiolar or perivascular infiltrate (untreated, n=9; silibinin, n=8; baricitinib, n=9). **(C)** Representative IF staining of lung-infiltrating neutrophils (left panel) and macrophages (right panel) in vFLIP mice. Scale bar, 20 µm. Cells were stained for DAPI (blue) and NE (red, left panel) or F4/80 (green, right panel). **(D)** tSNE representation of neutrophil subsets across untreated (3,741) mice and treated with silibinin (2,520) and baricitinib (2,694) colored according to cluster analysis (top). Stacked bar plots representing neutrophil clusters proportions across conditions. tSNE representation of monocytes/macrophages subsets across untreated (467) mice and treated with silibinin (342) and baricitinib (769) colored according to cluster analysis (bottom). Stacked bar plots representing monocytes/macrophages cluster proportions across conditions. **(E)** Functional assay performed at 1:3 ratio of PBMCs:$CD14^+$ cells using purified monocytes from COVID-19 patients (n = 5) treated with silibinin (200µm), baricitinib (200µm) or left untreated. Data are reported as mean ± S.E.M. *$p \leq 0.05$, **$p \leq 0.01$ and ***$p \leq 0.001$ by Mann–Whitney test (A,B,E).

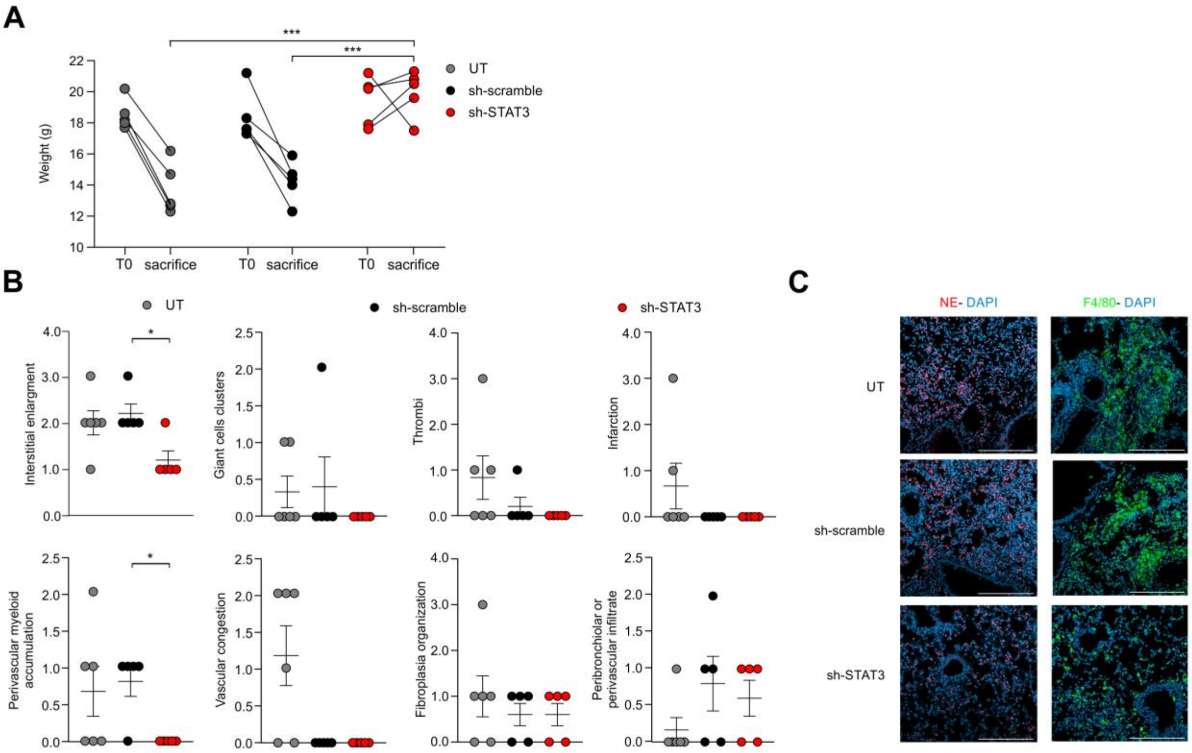
A
***
***
UT
sh-scramble
sh-STAT3
Weight (g)
T0
sacrifice
B
UT
sh-scramble
sh-STAT3
Interstitial enlargment
Giant cells clusters
Thrombi
Infarction
Perivascular myeloid accumulation
Vascular congestion
Fibroplasia organization
Peribronchiolar or perivascular infiltrate
C
NE- DAPI
F4/80- DAPI
UT
sh-scramble
sh-STAT3

**Fig. S6. Characterization of the in vivo STAT3-silencing effectiveness in vFLIP mice.**

**(A)** Body weights of vFLIP chimera mice before treatment (T0) or at the end of treatment (untreated, n=6; sh-scramble, n=5; sh-STAT3, n=5). **(B)** Dot plots of pathological score parameters: interstitial enlargement, giant cells clusters, thrombi, infarction, perivascular myeloid accumulation, vascular congestion, fibroplasia organization and peribronchiolar or perivascular infiltrate (untreated, n=6; sh-scramble, n=5; sh-STAT3, n=5). **(C)** Representative IF staining of lung-infiltrating neutrophils (left panel) and macrophages (right panel) in vFLIP mice. Scale bar, 20 µm. Cells were stained for DAPI (blue) and NE (red, left panel) or F4/80 (green, right panel). Data are reported as mean ± S.E.M. *$p \le 0.05$, **$p \le 0.01$ and ***$p \le 0.001$ by Mann–Whitney test.

### Table 1

**Table 1a. Main clinical data of patients for IHC analysisof lung autopsy .**

| | NRD | BP | COVID-19 |
|---|---|---|---|
| Parameters | N=4 | N=4 | N=23 |
| **Age (year), median (range)** | 64.5 (48-80) | 63 (44-82) | 82 (54-95) |
| **Gender, n (%)** | | | |
| Males | 2 (50%) | 4 (100%) | 11 (48%) |
| Females | 2 (50%) | | 12 (52%) |
| **Co-morbidity, n (%)** | | | |
| Absent | 0 | 1 (25%) | 3 (13%) |
| 1 | 1 (25%) | 2 (50%) | 8 (34.8%) |
| 2 | 0 | 1 (25%) | 8 (34.8%) |
| 3 | 3 (75%) | 0 | 4 (17.4%) |

**Table 1b. Main clinical data of COVID-19 patients for IHC analysis of lung autopsy.**

| | c-FLIP$^-$ CD68$^-$ pSTAT3$^-$ | c-FLIP$^+$ CD68$^+$ pSTAT3$^+$ |
|---|---|---|
| Days of hospitalization, median (range) | 25 (11-52); N=10 | 18 (0-32); N=13 |
| CRP (mg/L), median (range) | 75.79 (9-271.2); N=10 | 80.83 (5.92-242.5); N=12 |
| Pro-calcitonin (ng/mL), median (range) | 2.12 (0.14-9.45); N=9 | 0.81 (0.2-9.45); N=10 |

**Table 1c. Main clinical data of patients for molecular, phenotypic and functional data.**

| | Healthy controls | Mild patients | Severe patients |
|---|---|---|---|
| **Characteristics** | N=4 | N=11 | N=14 |
| Anagraphic | | | |
| Age, Yr: Median (IQR) | 66 (64-73) | 71 (63-86) | 70 (60-71) |
| Male, no. (%) | 3 (75) | 10 (90.9) | 6 (85.7) |
| **Clinical features at sampling** | | | |
| APACHE score, Median (IQR) | - | - | 25 (13.5-28) |
| SOFA score, Median (IQR) | - | - | 7 (4-8) |
| Score on ordinal scale (1-8), Median (IQR) | - | 4 (3-4) | 5 (3-7) |

**Supplementary materials and methods**

##### *Detection of cytokines and serology*

ProcartaPlex Mouse Cytokine & Chemokine Panel 1A (36 plex: IFN-γ; IL-12p70; IL-13; IL-1β; IL-2; IL-4; IL-5; IL-6; TNF-α; GM-CSF; IL-18; IL-10; IL-17A; IL-22; IL-23; IL-27; IL-9; GRO-α; IP-10; MCP-1; MCP-3; MIP-1α; MIP-1β; MIP-2; RANTES; Eotaxin; IFNα; IL-15/IL-15R; IL-28; IL-31; IL-1α; IL-3; G-CSF; LIF; ENA-78/CXCL5; M-CSF) (eBioscience, Thermo Fisher Scientific, Waltham, MA, USA) and Cytokine 25-Plex Human ProcartaPlex™ Panel 1B (25 plex: GM-CSF, IFN gamma, IL-1β, IL-2, IL-4, IL-5, IL-6, IL-12p70, IL-13, IL-18, TNF α, IL-9, IL-10, IL-17A (CTLA-8), IL-21, IL-22, IL-23, IL-27, IFN-α, IL-1 α, IL-1RA, IL-7, IL-15, IL-31, TNFβ (eBioscience, Thermo Fisher Scientific, Waltham, MA, USA) were performed according to manufacturer's instructions.

##### *Flow cytometry*

0.5-2x$10^6$ cells were washed in PBS and incubated with FcReceptor Blocking reagent CD16/32 (Biolegend) or FcReceptor Blocking reagent (Miltenyi Biotec) in staining buffer (2% FBS in PBS) for 10 min at 4 °C to saturate FcR. The following mAbs were then used for cell labelling: anti-mouse CD3 (17A2), CD45.1 (A20), CD11b (M1/70), B220 (RA3-6B2), CD45.2 (104), Ly6G (1A8), Ly6C (HK1.4), FOXP3 (NRRF-30), CD3ξ (145-2C11), CD25 (PC61.5), CD62L (MEL-14), CD8a (53-6.7), CD4 (RM4-5), LAG-3 (C9B7W), TIM-3 (B8.2C12), PD1 (29F.1A12), p-STAT3 (Ser727) (D4X3C), NK1.1 (PK136) or anti-mouse/human CD44 (IM7), p-STAT3 (pTyr705) (LUVNKLA) or human CD16 (3G8), CD3 (UCHT1), HLA-DR (L243), CD14 (MφP9), PD-L1 (MIH1) and Aqua LIVE/DEAD dye. All antibodies were purchased from the following companies: BD Biosciences (San Jose, CA, USA), eBiosciences (Thermo Fisher Scientific, Waltham, MA, USA), Biolegend (San Diego, CA, USA) and Cell Signaling Technologies (Danvers, MA, USA). Extracellular antigens were stained for 30 minutes at 4°C in staining buffer. For cytokines and transcriptional factor analysis, cells were fixed and permeabilized with Foxp3/Transcription Factor Staining Buffer Set (eBioscience) following manufacturer instructions. Intracellular antigens were stained for 1 hour in the appropriate 1x Perm/Wash buffer. Samples were acquired with a FACSCanto II (BD, Franklin Lakes, NJ, USA) and analyzed with FlowJo software (Treestar Inc.).

To determine the intracellular levels of p-STAT3 (Tyr705) and p-STAT3 (Ser727), after surface markers staining, cells were fixed with 2% paraformaldehyde (Sigma-Aldrich) and permeabilized with 90% cold methanol. For the intracellular staining, p-STAT3 (pTyr705) (clone LUVNKLA) antibody was used. All steps were performed in ice.

FLIP protein expression was evaluated by flow cytometry by indirect amplification on intracellular signal. In details, after surface markers staining, $1 \times 10^6$ PBMCs were fixed and permeabilized with Foxp3/Transcription Factor Staining Buffer Set (eBioscience). Before the intracellular staining, cells were incubated with FcReceptor Blocking reagent

(Miltenyi Biotec) for 10 min at RT in the appropriate 1x Perm/Wash buffer. Rabbit anti-FLIP antibody (D5J1E; 1:100; Cell Signaling Technologies) was added for 2 h at 4 °C. Signal was amplified with a secondary anti-rabbit IgG (H+L) (#8885, Cell Signaling Technology) for 30 minutes at 4 °C.

##### *Human cell isolation and functional assay*

Cells were isolated from EDTA-treated tubes (BD Biosciences, NJ, USA) and freshly separated by Ficoll-Hypaque (GE Healthcare) gradient centrifugation. PBMCs were counted and the monocyte fraction ($CD14^+$) was further isolated by CD14-microbeads (Miltenyi Biotec), following manufacturer's instructions. The purity of the $CD14^+$ fraction was evaluated by flow cytometry analysis. Samples with a purity greater than 95% were assessed for their suppressive capacity. $CD14^+$ cells were collected and cultured with 1 µM CellTrace (Thermo Fisher Scientific) labeled PBMCs, stimulated with coated anti-CD3 (OKT-3) and soluble anti-CD28 (28.2) for 4 days in 37°C and 8% $CO_2$ incubator. For the cells a ratio of 3:1 (target:effector) was used. At the end of the culture, cells were stained with anti-CD3 (UCHT1) and CellTrace signal of lymphocytes. Samples were acquired with FACS-Canto II (BD, Franklin Lakes, NJ, U.S.A.) using TruCountTM tubes (BD, Franklin Lakes, NJ, USA) to determine the absolute cell number of $CD3^+$ cells in the samples. Data were analyzed by FlowJo software (Tree Star, Inc. Ashland, OR, USA).

Cytokines released by patients' monocytes were quantified by Human ProcartaPlex™ Panel 1 multiplex. Samples with a purity greater than 95% were assessed for their cytokine production. Briefly, $5 \times 10^5$ $CD14^+$ cells were plated in 24-well plates for 12 hours. At the end of the incubation, viability was evaluated by flow cytometry.

##### *Immunofluorescence (IF) and immunohistochemistry (IHC)*

To determinate the presence of pSTAT3 Tyr705 and c-FLIP in human samples, $CD14^+$ cells were plated on coverslips (ibidi GmbH; Cat#80826), fixed in 4% formaldehyde for 10 minutes at RT, and blocked with 0.1% Triton X-100 in PBS 1x for 10 minutes at RT. To detect the intracellular signal, permeabilization was performed in 0.1% Triton X-100 in PBS 1x and 20% of normal goat serum (Vector Laboratories) for 2 hours at RT. Primary mAbs rabbit anti-FLIP antibody (D5J1E; 1:100; Cell Signaling Technologies) and mouse anti-pSTAT3 (Try705) (LUVNKLA; 1:50; Invitrogen) were diluted in PBS1x supplemented with 0,05% Tween 20 (Biorad, Cat#1706531) solution over-night at 4°C. Signal was amplified with secondary antibodies goat anti-rabbit IgG Alexa Fluor 488 (1:1000; CAT#A11034; Invitrogen) and donkey anti-mouse IgG Alexa Fluor 647 (1:1000; Cat#A-31571 Invitrogen) in 0.1% Triton X-100 in PBS 1x and 20% of normal goat serum for 1 hour at RT followed by nuclei staining with Hoechst 33342 (H1399; 1:500; Invitrogen) in PBS 1x for 10 minutes at RT.

Tissues were fixed in 10% neutral buffered formalin and embedded in paraffin; after embedding, 5µm thick sections were cut and stained with Hematoxylin and Eosin (Bio-Optica, Italy) for histological examination. For immunohistochemical and immunofluorescence analysis of samples, slides were deparaffinized, serially rehydrated and, after the appropriate antigen retrieval procedure, incubated with the following primary antibodies: rabbit anti-mouse pSTAT3 antibody (#9145, Cell Signaling), rat anti-mouse B220 antibody (550286, BD Pharmigen), mouse anti-human CD68 antibody (M0814, Dako), mouse anti-Human/Mouse/Rat FLIP antibody (MAB8430, R&D), rabbit anti-mouse CD3 antibody (ab16669, Abcam), rabbit anti-mouse F4/80 antibody (#70076, Cell Signaling), rabbit anti-mouse CD62P antibody (ab255822, Abcam), rabbit anti-mouse Neutrophil Elastase antibody (ab68672, Abcam), rabbit anti-mouse CD4 antibody (#25229, Cell Signaling) and rat anti-mouse Foxp3 antibody (14-5773-82, eBioscience), followed by the appropriate secondary antibodies. Immunostainings were developed with streptavidin peroxidase methods and the DAB Chromogen system (Dako). After chromogen incubation, slides were counterstained in Hematoxylin (Bio-Optica) and images were acquired by Leica DMRD optical microscope (Leica). For immunofluorescence, immunostainings were developed using TSA Plus Cyanine 3, TSA Plus Cyanine 5 or TSA Plus Fluorescein Systems (NEL744001KT, NEL745001KT or NEL741001KT respectively, Akoya Biosciences), goat anti-rabbit Alexa Fluor 488 (A11008, ThermoFisher), goat anti-rat Alexa Fluor 546 (A11081, ThermoFisher), goat anti-mouse Alexa Fluor Plus 647 (A32728, ThermoFisher) and nuclei were stained with Dapi (Sigma). Images were acquired by Zeiss LSM800 confocal microscope.

For histological assessment of collagen deposition, trichrome staining was performed using the Masson Trichrome with Aniline Blue Staining Kit (04-010802, Bio-Optica). Pathological score was independently evaluated by two pathologists in double-blind using the standard guideline previously published(*87*).

The percentage of CD3, B220 or F4-80 positive cells was evaluated on digital images of total reconstructed spleen section (5-10 X 50 microscopic fields per sample); clear brown positive cells were selected with the Magic Wand Tool of Adobe Photoshop. For each spleen, the number of positive cell pixels indicated in the histogram window was reported as % on the number of total spleen area (expressed in pixel). The number of Foxp3 positive cells was evaluated on digital images of immunofluorescence section as percentage on CD4 positive cells.

***Preparation of cell suspensions from organs***

Mice were euthanized by $CO_2$ inhalation. For lung flow cytometry analysis, mice were immediately perfused with 20 mL ice-cold PBS. Organs were harvested and processed as follows. Spleens were mechanically disaggregated and filtered (Corning Inc). Lungs were cut in small pieces with scissors, enzymatically digested at 37°C for 45 minutes with a solution containing collagenase IV (1mg/ml), hyaluronidase (0.1mg/ml) and DNase (4.5mg/ml) (Sigma-Aldrich). For bone marrow, tibias and femurs were flushed in RPMI 1640 (Euroclone) supplemented with 10% heat-inactivated FBS (Superior, Merck), 2mM L-glutamine, 10mM HEPES, 1mM sodium pyruvate, 150U/mL streptomycin, 200U/mL penicillin/streptomycin (all from Euroclone). Cells were then collected, filtered and red blood cells were lysed at RT for 5 minutes with ACK Lysing Buffer (Lonza). Peripheral blood was washed with PBS and red blood cells were lysed twice at RT for 10 minutes. Single-cell suspensions were then analysed by flow cytometry.

##### *Single-cell RNA sequencing (scRNA-seq)*

Lung pooled from 3 untreated-chimera, 3 silibinin-treated, 3 baricitinib-treated and 3 vFLIP-tg mice were digested as described in "Preparation of cell suspensions from organs" section. Single cell suspension ($10^4$ cells) was loaded on a GemCode Single Cell Instrument (10x Chromium System) to generate single cell GEMs.

*Single-cell RNA-seq data preprocessing*

Raw bcl files were demultiplexed using bcl2fastq v2.20 from Illumina and processed using 10x Cell Ranger v3.1.0. In particular, 'cellranger count' command with parameter '--expect-cells=4000' was used to quantify reads mapped to mouse (mm10) and human (GRCh38) genomes and to obtain the unique molecular identifier (UMI) count tables. Human scRNA-seq data from bronchoalveolar lavage fluids (BALs) was obtained from the Gene Expression Omnibus (GEO) under accession GSE157344.

*Single-cell RNA-seq data quality control*

The quality control steps before data integration were performed individually for each sample using the R package 'Seurat' (*88*) (*89*) (*90*) and Scrublet(*91*) for removing putative doublets. In particular, to retain high quality transcriptomes, the cells were filtered according to the following parameters: percentage of mitochondrial counts, minimum number of expressed genes, min./max. number of UMIs and doublet score. For both mouse and human only cells with less than 20% of total counts explained by mitochondrial genes were maintained. Scrublet thresholds were chosen seeing at the histograms of observed transcriptomes and simulated doublets as recommended by the author guidelines (https://github.com/AllonKleinLab/scrublet/blob/master/examples/scrublet_basics.ipynb). The other filtering thresholds were chosen looking at the distribution of the data in order to remove cells with a potential outlier behaviour. The complete list of filtering thresholds is summarized in the following table:

| **Sample** | **Min. number of expressed genes** | **Min. number of UMIs** | **Max. number of UMIs** | **Doublets threshold Scrublet** |
|---|---|---|---|---|
| WT 1 | 250 | 1,000 | 60,000 | 0.38 |
| WT 2 | 250 | 1,000 | 60,000 | 0.399 |

| | | | | |
|---|---|---|---|---|
| vFLIP 1 | 250 | 1,000 | 75,000 | 0.38 |
| vFLIP 2 | 250 | 1,000 | 75,000 | 0.52 |
| Chimera UT | 250 | 1,000 | 75,000 | 0.46 |
| Chimera Silibinin | 250 | 1,000 | 75,000 | 0.5 |
| Chimera Baricitinib | 250 | 1,000 | 75,000 | 0.38 |
| BAL3 | 200 | 500 | 40,000 | - |
| BAL7 | 200 | 500 | 40,000 | 0.689 |
| BAL10 | 200 | 500 | 100,000 | 0.4 |
| BAL29 | 200 | 500 | 60,000 | 0.627 |
| BAL18 | 200 | 500 | 40,000 | 0.661 |
| BAL24 | 200 | 500 | 100,000 | - |
| BAL25 | 200 | 500 | 60,000 | - |

*Single-cell RNA-seq data integration*

Both for mouse and human cells all the samples were integrated using the standard Seurat v3 integration procedure (*90*). Before data integration, the original count matrices were normalized using log-normalization from the Seurat package with default parameters. Next, both for mouse and human the scRNA-seq data was integrated using the first 30 dimensions of canonical correlation analysis (CCA). After integration, count matrices were scaled regressing for the total number of genes, UMIs and percentage of mitochondrial gene expression. Next, principal component analysis (PCA) was performed on the top 2,000 most variable features obtained using the 'vst' procedure of Seurat. The top 20 principal components were used to execute t-distributed stochastic neighbor embedding (tSNE) algorithm and to project the cells into a 2-dimensional space.

*Cell type identification*

After data integration, cell type identification both for mouse and human was performed using multiple reference-based cell annotation and manual inspection. For mouse, SingleR(*92*) with gene expression profiles from Immunological Genome Project(*93*) and Mouse RNA-seq(*94*) was used in combination with scMCA(*95*). For human data, SingleR was executed using reference gene signatures from Blueprint/ENCODE(*96*), (*97*), Human primary cell atlas (*98*) and Monaco immune data(*99*). Prior to final cell classification, cell labels were simplified in order to discard ultra-rare (< 20 cells) cell annotations and harmonized to match corresponding main population labels across the different reference datasets. Final cell identity was obtained taking the classification determined by 2 out of 3 reference datasets and annotating as "Unclassified" the cells labeled differently with all the 3 datasets. Finally, manual inspection was performed to check the final annotations and to solve part of the unclassified cells looking at the expression of known marker genes through appropriate dot plots (Figure S4B and S4D). Overall, about 2% of mouse and human cells were labeled as Unclassified.

*Cluster analysis of neutrophils and monocytes/macrophages*

To have a better resolution in subset cluster analysis, neutrophils and monocytes/macrophages populations of both mouse and human were extracted from the dataset, re-integrated using a similar approach to above (see Single cell RNA-seq data integration), and finally clustered. For neutrophils, cells were integrated using Seurat with 30 CCA dimensions and clustered using 10 principal components for mouse and 12 for human, respectively, to construct a shared nearest neighbor graph (SNN). A resolution of 0.12 for mouse and 0.15 for human, respectively, was used to find significant clusters (≥ 2% of cells) through the SNN. Monocytes/macrophages were similarly integrated and clustered using 11 and 10 principal components for mouse and human, respectively, with a resolution of 0.15. For human samples, the parameter 'k.filter=80' was set to find the Seurat integration anchors. After integration and clustering, cells were projected into a 2-dimensional space using the tSNE algorithm.

*Mapping between human and mouse genes*

To map mouse to human gene symbols the ortholog table from the Mouse Genome Informatics (MGI, http://www.informatics.jax.org/downloads/reports/HMD_HumanPhenotype.rpt) was used keeping only the genes with a one-to-one mapping between the two species (Table S2).

*Gene set analysis*

Gene set analysis for scRNA-seq and bulk RNA-seq data was performed using 'fgsea' (*100*) that performs pre-ranked gene set enrichment analysis (GSEA,(*101*)) or with the 'gsva' R package that performs gene set variation analysis (GSVA,(*102*)). Gene level statistics for scRNA-seq data used as input for GSEA were calculated using the Seurat function 'FindMarkers' with the Wilcoxon Rank Sum test with average log-fold change threshold of 0 for bulk-like analyses and 0.1 for cluster-level analyses. For the analysis of bulk RNA-seq of previous published Sars-CoV-2 ACE2-transgenic mice dataset (GSE154104), gene-level statistics were obtained through DESeq2 package(*103*). In particular, after removing duplicated gene symbols, differential expression analysis was performed to compare Sars-CoV-2 ACE2-transgenic mice 7 days post infection (dpi) and mock-infected mice (0 dpi) with log2-fold change different from 0. The 50 hallmark gene sets from MSigDB(*104*) were used as input both for GSEA and GSVA. For GSEA, only up- or down-regulated gene sets with adjusted p-value < 0.05 were considered statistically significant.

*Reference-based classification of human BAL neutrophils*

SingleR was used to label the cells of the neutrophil clusters in the human BALs using as reference the gene expression of vFLIP mice neutrophil clusters. In particular, a bulk-like mouse reference was created, taking the log-normalized average expression of each gene in each cluster, and giving it to SingleR for reference-based classification after mapping mouse genes to human gene symbols via orthology (see Mapping between human and mouse genes). SingleR was executed taking the top 100 differential expressed genes between cluster comparisons specifying the parameter 'de.n' (Table S2).

*Data visualization*

In general, the bioinformatics figures were obtained using functions of the R packages Seurat and ggplot2(*105*).
